# Exploring the Role of Chat GPT in patient care (diagnosis and Treatment) and medical research: A Systematic Review

**DOI:** 10.1101/2023.06.13.23291311

**Authors:** Ravindra Kumar Garg, Vijeth L Urs, Akshya Anand Agrawal, Sarvesh Kumar Chaudhary, Vimal Paliwal, Sujita Kumar Kar

**Affiliations:** Department of Neurology, King George’s Medical University, Lucknow, India: 226003; Department of Surgery, King George’s Medical University, Lucknow, India: 226003; Department of Neurology, Sanjay Gandhi Institute of Medical Sciences, Lucknow, India: 226001; Department of Psychiatry, King George’s Medical University, Lucknow, India: 226003

**Author notes:** **Correspondence**: Ravindra Kumar Garg, Department of Neurology, King George’s Medical University, Lucknow, India: 226003.

**Keywords:** Artificial intelligence, Plagiarism, Authorship, Scholarly publishing

## Abstract

**Background:** ChatGPT(Chat Generative Pre-trained Transformer) is an artificial intelligence (AI) based on a natural language processing tool developed by OpenAI (California, USA). This systematic review examines the potential of Chat GPT in diagnosing and treating patients and its contributions to medical research.

**Methods:** In order to locate articles on ChatGPT’s use in clinical practise and medical research, this systematic review used PRISMA standards and conducted database searches across several sources. Selected records were analysed using ChatGPT, which also produced a summary for each article. The resultant word document was transformed to a PDF and handled using ChatPDF. The review looked at topics pertaining to scholarly publishing, clinical practise, and medical research.

**Results:** We reviewed 118 publications. There are difficulties and moral conundrums associated with using ChatGPT in therapeutic settings and medical research. Patient inquiries, note writing, decision-making, trial enrolment, data management, decision support, research support, and patient education are all things that ChatGPT can help with. However, the solutions it provides are frequently inadequate and inconsistent, presenting issues with its originality, privacy, accuracy, bias, and legality. When utilising ChatGPT for academic writings, there are issues with prejudice and plagiarism, and because it lacks human-like characteristics, its authority as an author is called into question.

**Conclusions:** ChatGPT has limitations when used in research and healthcare. Even while it aids in patient treatment, concerns regarding accuracy, authorship, and bias arise. Currently, ChatGPT can serve as a “clinical assistant” and be a huge assistance with research and scholarly writing.

## Introduction

ChatGPT(Chat Generative Pre-trained Transformer) is an artificial intelligence (AI) based on a natural language processing tool developed by OpenAI (California, USA). ChatGPT is chat boat based technology. A chatbot is in fact a type of software creates text akin to human-like conversation. ChatGPT has the capacity to respond to follow-up questions, recognise errors, debunk unfounded theories, and turn down inappropriate requests. Large Language Models (LLMs), which are frequently abbreviated as LLMs, are extremely complex deep-learning programmes that are capable of comprehending and producing text in a manner that is strikingly comparable to that of humans. LLMs can recognise, summarise, translate, predict, and create text as well as other sorts of information by using the large knowledge base they have amassed from massive datasets.^1, 2^

The possible uses of ChatGPT in medicine is currently under intense investigation. ChatGPT is considered to have enormous capability in helping experts with clinical and laboratory diagnosis to planning and execution of medical research. Another significant use of ChatGPT in medical researchers is the creation of virtual assistants to physicians helping them in writing manuscripts in more efficient way. Usage of ChatGPT in medical writing is considered to have associated with several ethical and legal issues. Possible copyright violations, medical-legal issues, and the demand for openness in AI-generated content are a few of these.^3–5^

In this systematic review we aimed to review published article and explore the potential of ChatGPT in facilitating patient care, medical research and medical writing. We will also focus on ethical issues associated with usage of ChatGPT.

## Methods

We performed a systematic review of published articles on ChatGPT. The protocol of the systematic review was registered with PROSPERO (PROSPERO 2023 CRD42023415845).^6^ Our systematic review was conducted following the Preferred Reporting Items for Systematic Reviews and Meta-Analyses (PRISMA) guidelines.

## Search strategy

We searched four databases, PubMed, Scopus, Embase, and Google Scholar. Our search was aimed at identifying all kinds of articles on ChatGPT and its application in medical research, scholarly and clinical practice, published till 24 May 2023. Articles related to medical education was not considered. The search item that we used was “ChatGPT”. We reviewed all kinds of publications including original articles, reviews, editorial/ commentaries and even letter to the editor describing ChatGPT.

## Data extraction

The selection of the papers that were published was done in two steps. Two reviewers (RK and VKP) reviewed the titles and abstracts in the initial phase. Two reviewers (VU and SKC) then examined the entire texts of the chosen papers to determine their eligibility. A third author (SK) settled any differences that arose between the two authors. Two reviewers (RK and VKP) assessed the information available in the included publication for the suitability of the article to be included in the review. Any disagreement between them was resolved by mutual agreement. If a dispute persisted, it was resolved via consultation with a third reviewer (SK).

EndNote 20 web tool (Clarivate Analytics) was used to handle duplicate records. This process was carried out by two reviewers independently (RK and VKP). Any issue that arose was resolved with a discussion with another reviewer. The number of retrieved and assessed records at each stage was provided in the form of a PRISMA flow chart. EndNote 20 (Clarivate Analytics) was used to make a PRISMA flow chart.

## Quality assessment

Quality assessment was not done.

## Data analysis

ChatGPT was extensively used for analysing the selected records and writing this manuscript. A table was made with six columns (First author/sole author, country of origin, status of peer review (peer-reviewed or preprint), title of the paper and short point wise summary of full text. Short point wise summary of full text of each and every article was created with the help of ChatGPT. The voluminous word file was then converted to a pdf file and was processed with the sister software “ ChatPDF” (OpenAI, California, USA available at https://www.chatpdf.com/). Following questions were asked from ChatPDF.

1. What are potential role of ChatGPT in medical writing and research?
2. What could be the role of ChatGPT in clinical practice?
3. What are ethical issues associated with paper writing?
4. Can ChatGPT be an author?
5. Can ChatGPT write text in good English and free of plagiarism?
6. Role of ChatGPT so far in neurological disorders related clinical practice and research.
7. Effectiveness and efficiency of ChatGPT in medical research and clinical settings
8. Potential benefits and limitations of ChatGPT in medical research and clinical applications
9. The ethical implications of using ChatGPT in medical research and clinical practice
10. Identify the gaps in the current research on ChatGPT and suggest areas for further investigation.
11. Provide insights into the potential future applications of ChatGPT in medical research and clinical practice
12. Recommendations for researchers, clinicians, and policymakers on the use of ChatGPT in medical research and clinical practice

All the responses were compiled in a word file.

## Results

Our data collection followed PRISMA guidelines. (Additional file 1: PRISMA checklist) The PRISMA flowchart for our systematic review is shown in Figure 1. We reviewed 118 publications. ChatGPT related publications are available from across the globe. There were 33 original articles and rest were commentary/ editorial, review articles, research letters or letter to the editors. Out of 118 articles, 18 articles were available as preprint only. Summaries of 118 articles and answers to 12 questions have been provided in form of tables. (Table-1 and Table-2)

**Figure 1:**
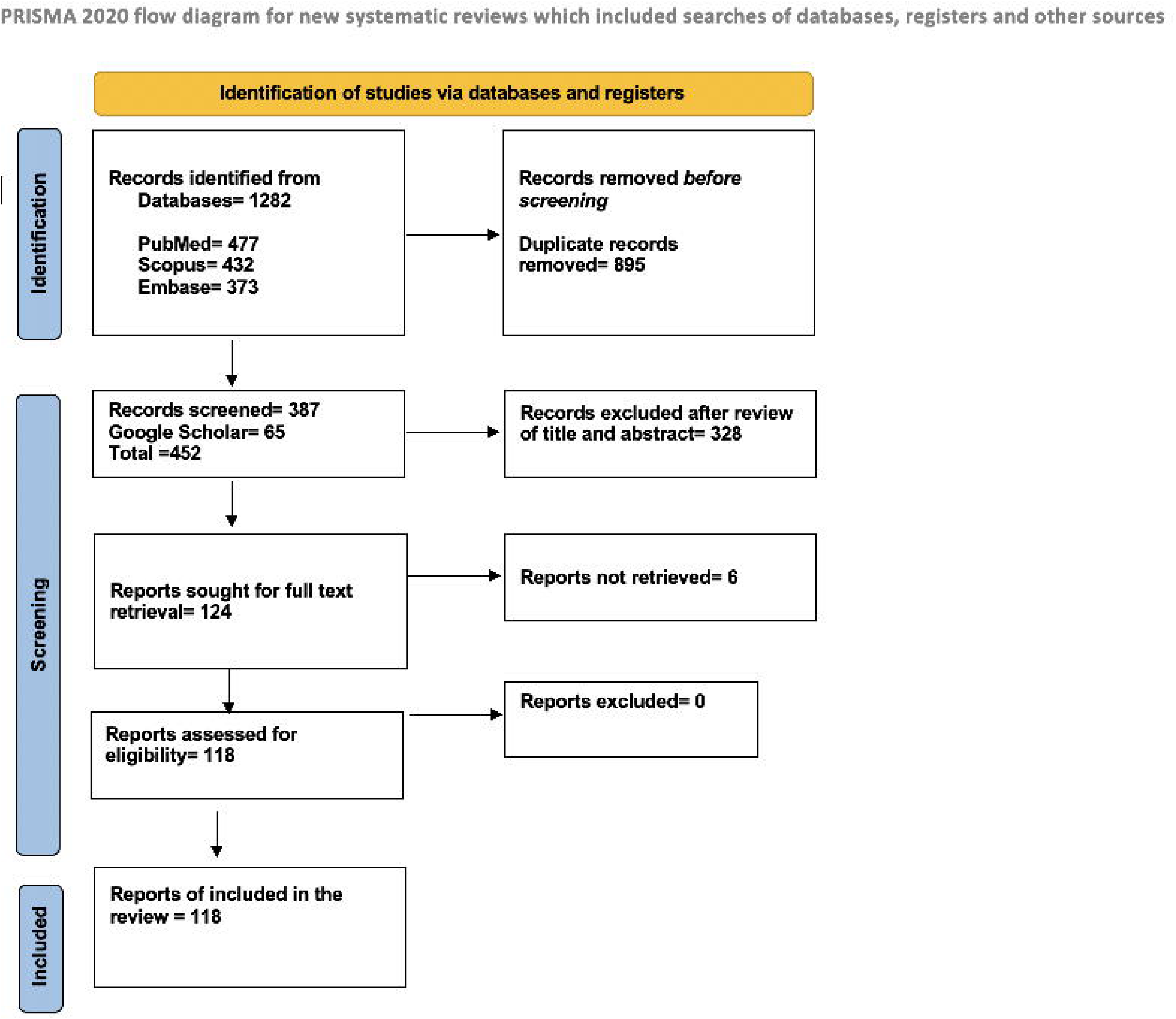
The study’s PRISMA flow diagram shows how articles are selected for this systematic review.

**Table-1:** Summaries of full text of 118 selected article. Summaries were created with the help of ChatGPT. PROMPT, that given was “Provide point wise summary” .

| Reference | Country | Type of publication | Status of peer review | Title of the paper | Main findings of the paper |
| --- | --- | --- | --- | --- | --- |
| Ali and Djalilian 2023 | India | Editorial | Peer reviewed | Readership awareness series–paper 4: Chatbots and ChatGPT-ethical considerations in scientific publications | The question of whether a chatbot like ChatGPT can be regarded as an author gives rise to ethical dilemmas concerning accountability for the content produced and the accuracy of source attributions. |
| Ali et al 2023 | United Kingdom | Commentary | Peer reviewed | Using ChatGPT to write patient clinic letters | CHATBOTS, such as ChatGPT, were employed by researchers to generate a variety of clinical communication scenarios that were designed to be comprehensible for individuals at or below a sixth-grade level (approximately 11-12 years of age). |
| Alser and Waisberg 2023 | Egypt | Commentary | Peer reviewed | Concerns with the Usage of ChatGPT in Academia and Medicine: A Viewpoint | ChatGPT, a language model, was credited authorship in four medical papers. It met one of four ICMJE authorship criteria by contributing to writing parts of the papers. Plagiarism was found in its contributions. Unclear sources of information and bias due to tuning also limit its use in scientific contexts. |
| Anderson et al 2003 | Germany and other European countries | Editorial | Peer reviewed | AI did not write this manuscript, or did it? Can we trick the AI text detector into generated texts? The potential future of ChatGPT and AI in Sports & Exercise Medicine manuscript generation | The ability of AI to produce academic papers has been tested and found wanting. The papers produced by AI lacked fresh ideas, in-depth knowledge of the subject, and precise referencing. There were problems with equality, accuracy, poor AI detection, unethical content creation, and plagiarism. To maintain the quality of scientific literature, the application of AI tools needs to be carefully studied. AI misuse can be avoided with the aid of current plagiarism-detection tools and possible cutting-edge software for AI-generated text detection. |
| Arun Babu and Sharmila 2023 | India | Letter to the editor | Peer reviewed | Using artificial intelligence chatbots like 'ChatGPT' to draft articles for medical journals - Advantages, limitations, ethical concerns and way forward | The use of AI-generated content in medical journals raises ethical concerns regarding accuracy, bias, authorship, and disclosure, necessitating new guidelines and standards for publication integrity. |
| Asch 2023 | USA | Commentary Interview and chat with | Peer reviewed | An Interview with ChatGPT About Health Care | ChatGPT has potential in healthcare, assisting with tasks like virtual assistance, documentation, research, education, and patient engagement. Concerns such as privacy, bias, regulation, and ethics must be addressed. It can both improve and reduce equity. Advancements should focus on data quality, NLP, integration, |
|  |  | ChatGPT |  |  | security, explainability, and ethics. It enhances, not replaces, healthcare professionals. |
| Athaluri et al 2023 | India | Original | Peer reviewed | Exploring the Boundaries of Reality: Investigating the Phenomenon of Artificial Intelligence Hallucination in Scientific Writing Through ChatGPT References | This study investigated AI hallucination in research proposals drafted by ChatGPT. Analysis of 178 references revealed limitations in generating reliable references, including the absence of DOIs and inaccessible articles. The findings emphasize the need for improvements in training and caution when relying solely on ChatGPT's generated references. |
| Balas and Ing 2023 | Canada | Original | Peer reviewed | Conversational AI Models for ophthalmic diagnosis: Comparison of ChatGPT and the Isabel Pro Differential Diagnosis Generator | When it came to accurately recognising the diagnosis in 9 out of 10 cases and including it in all differential diagnosis lists, ChatGPT performed better than Isabel Pro at diagnosing ocular illnesses. ChatGPT has potential for primary care physicians. |
| Barker and Rutka 2023 | Editor-in-Chief, Journal of Neurosurgery | Editorial | Peer reviewed | Generative artificial intelligence, chatbots, and the Journal of Neurosurgery Publishing Group | In a comparative study, ChatGPT answered 60.2% of neurosurgical questions correctly, while average human users scored 69%. However, caution is necessary as chatbot outputs may contain biases, inaccuracies, and even fabrications. Trusting chatbot-generated text without verification can lead to misleading information. |
| Bauchner 2023 | former editor-in-chief of JAMA USA | Original | Peer reviewed | ChatGPT: Not An Author, But A Tool | The ICMJE's requirements for authorship are not currently met by ChatGPT. However, it might be utilised by researchers as a tool to design early publications, possibly complying to reporting standards and offering a more objective viewpoint. Additionally encouraging diversity, it might help non-native English speakers prepare papers for publication. ChatGPT can be recognised in articles along with other pertinent information as a method to improve scientific communication. |
| Baumgartner 2023 | Austria | Commentary | Peer reviewed | The potential impact of ChatGPT in clinical and translational medicine | The utilization of ChatGPT in clinical and translational medicine has the potential to bring about a transformative impact, enhancing patient engagement, reducing healthcare providers' workload, and delivering current information. Nonetheless, it is imperative to confront challenges such as data privacy risks, limitations in accuracy, and potential bias originating from training data. Ongoing research and development efforts are crucial to ensure the safe and effective implementation of ChatGPT within the healthcare domain. |
| Benoit 2023 | Canada | Original | Preprint | ChatGPT for Clinical Vignette Generation, Revision, and Evaluation | This study examined the capabilities of ChatGPT in generating, rewriting, and evaluating clinical vignettes. ChatGPT demonstrated the ability to follow instructions, generate varied contexts, and match patient demographics. However, there were limitations and caveats, requiring monitoring and review. The findings highlight the potential of ChatGPT with appropriate oversight. |
| Bhattacharya et al | India | Review | Peer | ChatGPT in Surgical | ChatGPT in anaesthesia and surgical care has potential for clinical decision- |
| 2023 |  |  | reviewed | Practice—a New Kid on the Block | making, preoperative education, medical record transcription, surgical planning, and medical education. However, limitations include potential misinformation, bias, reliance on outdated data, copyright concerns, technical issues, and lack of personalization. Proper evaluation, oversight, and addressing legal and ethical considerations are necessary for optimal use. |
| Biswas 2023 | USA | Commentary | Peer reviewed | ChatGPT and the future of medical writing | Article introduces AI and ChatGPT. |
| Boßelmann et al 2023 | USA | Commentary | Peer reviewed | Are AI language models such as ChatGPT ready to improve the care of individuals with epilepsy? | Artificial Intelligence (AI) can significantly improve epilepsy diagnosis and treatment. Examples of epilepsy-related AI research include EEG data analysis, MRI-based lesion detection, and Clinical Decision Support Systems. While AI language models like ChatGPT show promise in improving patient care, they require rigorous testing, validation, and ethical consideration, along with the expertise of medical professionals. |
| Brainard 2023 | USA | Commentary | Peer reviewed | Journals take up arms against AI-written text | Publishing directors and journal managers are concerned about the impact of AI-powered chatbots like ChatGPT on scholarly literature. These tools can produce accurate-sounding, AI-generated reports and scientific manuscripts, raising issues of accuracy, authorship, and potential flooding of AI-produced manuscripts. Publishers are formulating policies requiring disclosure of AI tool use and are exploring technology to detect synthetic text. |
| Cahan and Treutlein 2023 | Switzerland | Editorial | Peer reviewed | A conversation with ChatGPT on the role of computational systems biology in stem cell research | Computational biology and systems biology have made significant contributions to stem cell research by enabling the analysis of large datasets, identifying patterns and trends, optimizing stem cell culture conditions, and creating detailed models of stem cell behavior. Challenges include the need for extensive data collection and processing, but advancements in data standardization and accessibility have facilitated data reuse and meta-analyses. Integrating computational techniques with stem cell research holds great promise for advancing our understanding and applications of stem cells. |
| Nasrallah 2023 | Editor-in-Chief Current Psychiatry | Editorial Guest editorial generated by ChatGPT | Peer reviewed | A 'guest editorial' ... generated by ChatGPT? | The article discusses the introduction of ChatGPT, an AI program with potential societal impact. It highlights concerns about accuracy and references, tests ChatGPT's abilities, and reflects on the era of AI-generated articles. |
| Cascella et al 2023 | Italy | Review | Peer reviewed | Evaluating the feasibility of ChatGPT in healthcare: an analysis of multiple clinical and research scenarios | This study examines ChatGPT's possible uses in clinical and research settings, including supporting clinical practice, doing scientific work, preventing potential abuse, and debating public health issues. The study highlights ChatGPT's shortcomings and emphasises the need for careful use and further development to avoid dangers, such as disinformation and misuse, even if it shows promise in a number of areas. |
| Chen et al 2023 | USA | Original | Preprint | The utility of ChatGPT for cancer treatment information | This study found that while ChatGPT correctly aligned with National Comprehensive Cancer Network (NCCN) guidelines in 98% of cases, it also provided partially non-concordant advice in 34.3% of them. Therefore, it concluded that ChatGPT is not entirely reliable for cancer treatment advice, urging patients and clinicians to recognize its limitations. |
| Cheng et al 2023 | China | Letter to the editor | Peer-reviewed | ChatGPT/GPT-4: Enabling a new era of surgical oncology | The potential application of ChatGPT/GPT-4 in surgical oncology, from aiding in clinical trial design, case management, data analysis, to preoperative preparations and intraoperative processes, is discussed in this study. Despite potential challenges and ethical concerns, the authors advocate for responsibly harnessing this AI technology to improve surgical oncology outcomes. |
| Chervenak et al 2023 | USA | Original | Peer-reviewed | The promise and peril of using a large language model to obtain clinical information: ChatGPT performs strongly as a fertility counseling tool with limitations | ChatGPT, tested against established fertility sources, provided responses comparable in quality to the CDC's FAQs and scored highly on validated fertility knowledge surveys. Although it effectively reproduced missing facts from the American Society for Reproductive Medicine's committee opinion, its clinical utility might be limited by occasional incorrect information and lack of source citation. |
| Cifarelli and Sheehan 2023 | USA | Commentary | Peer-reviewed | Large language model artificial intelligence: the current state and future of ChatGPT in neuro-oncology publishing | However, ethical considerations arise when utilizing ChatGPT in scholarly publishing. These encompass potential bias in training data and the perpetuation of prejudice, concerns about authorship, copyright, and plagiarism, implications for citation practices and the "Matthew Effect," and impacts on academic job expectations, tenure, and promotion.<br>Addressing ownership of generated content and adherence to copyright laws is crucial, necessitating careful source attribution. Plagiarism is a concern, but proper citation practices can help mitigate it.<br>ChatGPT can streamline citation processes and assist researchers in identifying and formatting citations accurately. However, relying solely on automated tools may overlook important literature and raise questions about the value of human expertise.<br>Academic institutions and publishers can implement measures such as anti-ChatGPT software, encourage creative and innovative research, and reconsider tenure evaluation criteria to tackle the challenges posed by ChatGPT.<br>Thoughtful consideration of ethical concerns is essential to ensure responsible and ethical use of ChatGPT in academia and scholarly publishing. Further research is warranted to explore its implications and limitations. |
| Corsello and Santangelo 2023 | Italy | Review | Peer-reviewed | May Artificial Intelligence Influence Future Pediatric Research?—The Case of ChatGPT | This interview with ChatGPT explores the transformative potential of AI in pediatric research, highlighting advantages such as improved clinical decision-making, enhanced education, faster drug development, and better research outcomes. It also probes into potential challenges like bias, safety issues, overreliance on technology, and ethical concerns. It underscores the importance of careful consideration of these technologies' implications and usage, ensuring they are responsibly leveraged for beneficial outcomes. |
| D'Amico et al 2023 | USA | Editorial | Peer-reviewed | I asked a ChatGPT to write an editorial about how we can incorporate chatbots into neurosurgical research and patient care... | This editorial discusses the potential and challenges of incorporating AI chatbots in neurosurgery for data collection, patient care, and communication. However, issues like potential misinformation, privacy, ethical concerns, bias, legal liabilities, content validity, and effectiveness need addressing. It emphasizes human responsibility in verifying machine-generated content for moral and ethical standards. Neurosurgery should lead in responsibly integrating these AI technologies. |
| Darkhabani et al 2023 | Turkey | Review | Peer-reviewed | ChatGPT and autoimmunity - A new weapon in the battlefield of knowledge | This paper explores the role of AI language model ChatGPT in clinical medicine, specifically autoimmunity, discussing its capabilities, limitations, and potential cyber risks. It highlights the importance of continuous evaluation, as ChatGPT and similar technologies evolve rapidly, emphasizing the need for healthcare professionals to stay abreast of these developments. |
| Dave 2023 | UK | Commentary | Peer-reviewed | Plagiarism software now able to detect students using ChatGPT | Turnitin, the academic integrity firm used by 98% of UK universities, can now detect plagiarism in content generated by artificial intelligence systems like ChatGPT. This development aims to maintain academic integrity in the face of advancements in AI technology, preventing dishonest use of AI-generated work. |
| Dave et al 2023 | UK | Review | Peer-reviewed | ChatGPT in medicine: an overview of its applications, advantages, limitations, future prospects, and ethical considerations | ChatGPT can be a valuable tool for medical research and patient care, providing support in data analysis and diagnosis generation. However, it should supplement, not replace, human expertise. Ethical considerations, like data privacy and accuracy of recommendations, need to be addressed. Future prospects include integration with other AI technologies, prioritizing a balanced, cautious, and ethically mindful approach. |
| Day et al 2023 | Canada | Original article | Peer-reviewed | A Preliminary Investigation of Fake Peer-Reviewed Citations and References Generated by ChatGPT | ChatGPT provide citations as well although recent scrutiny revealed that these references are often incorrect or non-existent, according to a study. Despite the impressive capabilities of the chatbot, the presence of fake references and citations is a concern for both academic research and student academic integrity. While these inaccuracies present a problem for those who might use the bot for sourcing, they could serve as a useful tool for identifying academic misconduct or as prompts for further research. Despite these issues, the chatbot has been utilized effectively for generating course materials in certain subject areas, suggesting potential for its continued use in education. However, the tool requires subject expertise and its use should be approached with caution. |
| de Oliveira and Ballesterro 2023 | Brazil | Editorial | Peer-reviewed | The future of Pediatric Neurosurgery and ChatGPT: an editor's perspective | The use of AI like ChatGPT in medical writing is revolutionary, but has ethical implications and potential for errors. AI's usage should be transparent, and authors should ensure their publications' scientific integrity. |
| De Vito 2023 | Argentina | Editorial | Peer-reviewed | Artificial intelligence and chatGPT. Would you read an artificial author? | This editorial discusses the implications of using AI, particularly OpenAI's GPT-3, in scientific and academic writing. While acknowledging the benefits of AI tools in producing coherent text, it raises concerns about authenticity, potential misuse, and the need for transparency. It also stresses the importance of human responsibility in ensuring the accuracy of scientific content and the ethical use of AI technology in academia. |
| Dergaa et al 2023 | Multiple | Original | Peer- | From human writing to | This study explores the benefits and potential threats of NLP technologies like |
|  | countries | article | reviewed | artificial intelligence generated text: examining the prospects and potential threats of ChatGPT in academic writing | ChatGPT in academic writing and research. While these tools can enhance efficiency, concerns arise about the authenticity and credibility of work. The study underscores the need for ethical considerations, transparency, and human intelligence in their use to maintain academic integrity. |
| Donato et al 2023 | Portugal | Editorial | Peer reviewed | The Transparency of Science with ChatGPT and the Emerging Artificial Intelligence Language Models: Where Should Medical Journals Stand? | This article discusses ethical implications of large language models (LLMs) like ChatGPT in scientific publishing. It highlights concerns about authorship, transparency, and integrity in utilizing AI tools. The need to declare AI's role and limit its misuse is emphasized. It also discusses future prospects of detecting AI-generated text and potential applications of LLMs in non-English language contexts. |
| Dunn et al 2023 | USA | Original article | Peer reviewed | Artificial intelligence-derived dermatology case reports are indistinguishable from those written by humans: A single-blinded observer study | This letter discusses a study comparing the quality and readability of dermatology-based case reports produced by human authors and AI models like ChatGPT. Findings suggest that AI-generated reports were often indistinguishable from human-created ones, though some limitations were noted. AI detection tools varied in effectiveness, raising ethical implications about the reliability, accuracy, and transparency of AI-generated content. The authors argue for the need to maintain integrity of AI-produced content, and for editorial teams to consider policies around the use of AI and Large Language Models. |
| Fatani 2023 | Saudi Arabia | Review | Peer reviewed | ChatGPT for Future Medical and Dental Research | This narrative review discusses the application and implications of AI language model, ChatGPT, in medical and dental research. ChatGPT can assist in writing scientific papers, summarizing data, and translating languages. While AI can enhance clinical workflow and help generate quick responses, it's cautioned that over-reliance could lead to papers lacking critical thinking. Ethical concerns such as plagiarism and data integrity are also highlighted. The review concludes that while ChatGPT can support research and potentially transform clinical medicine, there are limitations and potential risks that need to be carefully managed. |
| Galland 2023 | France | Editorial | Peer reviewed | Chatbots and internal medicine: Future opportunities and challenges | In November 2022, OpenAI® made headlines by releasing version 3.5 of its AI ChatGPT, a revolutionary conversational AI capable of generating text on any topic, including in the medical field. It was joined in 2023 by other advanced generative AIs such as Google's BardAI and image generators like Dall-E 2 and Mid Journey. The use cases in healthcare are increasing, and chatbots could be valuable tools for prevention, appointment scheduling, symptom information, administrative tasks, and more. However, concerns about data privacy, user safety, and accuracy of information arise with the use of chatbots in healthcare. The adoption of these technologies by patients may outpace the establishment of scientific consensus on their use. The article emphasizes the need for caution, evaluation, and defining the indications and limitations of their utilization in the medical field. |
| Gandhi | India | Letter to the | Peer | ChatGPT: roles and | The article discusses the advancements in AI, particularly ChatGPT, an advanced |
| Periaysamy et al 2023 |  | editor | reviewed | boundaries of the new artificial intelligence tool in medical education and health research - correspondence | language model developed by OpenAI. It highlights its potential in medical education and health research. ChatGPT can provide answers and information for medical students, assist researchers in writing and reviewing content, and even generate scientific abstracts. However, it also raises concerns about potential misuse, integrity of generated content, and ethical considerations regarding authorship in research papers involving AI. Proper guidelines and reporting standards are recommended to ensure responsible and transparent use of AI tools in the medical field. |
| Gao et al 2023 | USA | Original article | Peer-reviewed | Comparing scientific abstracts generated by ChatGPT to original abstracts using an artificial intelligence output detector, plagiarism detector, and blinded human reviewers | The article assesses ChatGPT's capacity to produce scientific abstracts, a sizable language model. Although neither was flawless, both human reviewers and AI output detectors were able to recognise some created abstractions. The created abstracts were in the same range as actual abstracts but contained made-up numbers. The study is aware of its limitations, including the sample size and the type of cues employed. The technology has the potential to be used in both ethical and immoral ways, such as to fabricate research or aid in research writing. The biases in training data are underlined, as is the requirement for more investigation into potential biases. |
| Goedde et al 2023 | Germany | Original article | Preprint | ChatGPT in medical literature-a concise review and SWOT analysis | This concise review focuses on ChatGPT (Chat Generative Pre-trained Transformer) and its role in medical literature. The study analyzed the literature published on ChatGPT from December 2022 to March 2023 and conducted a SWOT analysis. The findings highlight the strengths, weaknesses, opportunities, and threats associated with ChatGPT. The review emphasizes the need for further research, regulations, and policies to address the potential of ChatGPT in medical literature. |
| Gordijn and Have 2023 | Ireland | Editorial | Peer-reviewed | ChatGPT: evolution or revolution? | This editorial explores the implications of large language models (LLMs), specifically ChatGPT by OpenAI, on academic publishing and research. It questions whether the development of LLMs will necessitate new editorial policies to address the challenges they pose. Viewing ChatGPT as either evolutionary or revolutionary, the article suggests the utility of the AI model as a research tool may be limited due to deficiencies. It acknowledges the potential for LLMs to improve research but also highlights potential risks such as plagiarism and factual inaccuracies. Current editorial policies may already handle these issues, but further review and adjustments could be necessary. |
| Gottlieb et al 2023 | USA | Commentary | Peer-reviewed | ChatGPT and conversational artificial intelligence: Friend, foe, or future of research? | AI and machine learning are significantly impacting healthcare and research, enhancing precision medicine through big data analysis and personalized treatment plans.<br>Large Language Models (LLMs) like ChatGPT serve as conversational interfaces, aiding in patient communication and assisting researchers in content generation. Advantages include accelerating content creation, language translation, simplifying complex topics, synthesizing literature, creating abstracts, identifying limitations, and disseminating knowledge.<br>Potential harms encompass limitations in the extant knowledge of LLMs, potential misinterpretation of study methods/results, overconfidence in AI-generated content,<br>risk of plagiarism, inaccurate references, and biases.<br>Current trends indicate the use of AI for language editing and translation of academic papers, increasing global access.<br>Controversy persists regarding AI's authorship status in academic research; some argue that AI fails to meet authorship criteria as it can't be accountable for the work. A balance must be struck, leveraging AI benefits while ensuring ethical standards, accuracy, and acknowledging inherent limitations. |
| Graf and Bernardi 2023 | Germany | Editorial | Peer-reviewed | ChatGPT in Research: Balancing Ethics, Transparency and Advancement | ChatGPT can generate realistic text, propose research questions, design studies, analyze data, write and edit documents, among other tasks, offering valuable starting points or improvements for researchers' work.<br>The ethics and appropriateness of listing ChatGPT as a co-author in research papers has stirred debate in the scientific community.<br>Some argue that if ChatGPT contributes to the planning, execution or manuscript writing/editing of a study, it should be credited as a co-author.<br>However, editors-in-chief from major scientific journals disagree, arguing that authorship requires the capacity for accountability and consent, which ChatGPT, as an AI, lacks.<br>ChatGPT agrees with this sentiment, stating it doesn't possess the ability to consent to co-authorship, but can assist with writing and editing.<br>Despite these discussions, the AI's contributions are currently undetected by plagiarism checker software, raising concerns about originality and authorship in academic work.<br>A recent study showed that ChatGPT-generated abstracts were often indistinguishable from "original work," both by plagiarism checkers and human academic reviewers.<br>As AI continues to evolve rapidly, it poses significant questions for publishers, researchers and educators about how to best utilize, regulate and attribute its capabilities |
| Graham 2023 | UK | Commentary | Peer-reviewed | ChatGPT and other AI tools put students at risk of plagiarism allegations, MDU warns | As advanced AI technology becomes more accessible, the MDU (Medical Defence Union) is emphasizing the importance of students ensuring that their academic work remains "above suspicion."<br>Ellie Mein, a medicolegal adviser at MDU, cautions against taking shortcuts to meet deadlines and advises struggling students to seek support or extensions instead. |
|  |  |  |  |  | Plagiarism allegations can lead to fitness to practice investigations according to the guidelines set by the General Medical Council (GMC), which assesses whether students meet the required competency standards.<br>Universities have the authority to impose conditions, suspension, or expulsion on students found to be involved in plagiarism, even if it was unintentional.<br>When applying for full registration, students are obligated to disclose any fitness to practice investigations to the GMC.<br>Mein highlights that some students have been taken aback when contacted by the GMC about undisclosed investigations.<br>To help students avoid plagiarism in the "age of AI," the MDU has provided tips that address common errors such as "para-plagiarizing" and insufficient citation. |
| Gravel et al 2023 | Canada | Original article | Preprint | Learning to fake it: limited responses and fabricated references provided by ChatGPT for medical questions | The objective of this study was to assess the quality and suitability of responses and references provided by ChatGPT when addressing medical questions. The study selected 20 diverse medical questions derived from recent research articles published in high-impact factor medical journals. These questions were related to the main objectives of the articles or framed within a broader context to ensure a comprehensive range of references.<br>The study followed a specific methodology where the questions were posed to ChatGPT without imposing any word limit or constraint. After receiving the response, a follow-up query was made to request references, with only the first three references being considered for analysis. To ensure domain expertise, the corresponding authors of the selected articles were invited to rate the responses. The primary outcomes measured in the study were the appropriateness of the references and the quality of the responses.<br>The findings of the study revealed that the majority of references provided by ChatGPT were fabricated. These references appeared credible, featuring authors with previous publications or affiliations with reputable organizations. However, 69% of the references were found to be nonexistent. Among the 59 references included in the analysis, only 18 were authentic, although they contained minor or major citation errors. The remaining references were completely fictional. Additionally, the study reported that 95% of the references listed authors with prior publications or had affiliations with recognized organizations.<br>Among the participating corresponding authors, seventeen agreed to evaluate the responses, assigning a median score of 60% for the quality of the answers. Notably, five responses were identified by the raters to contain major factual errors. |
|  |  |  |  |  | To summarize, the study revealed that the references provided by ChatGPT were predominantly fabricated, and the quality of the responses varied, including instances of major factual errors. These findings raise concerns regarding the reliability and accuracy of the references and responses generated by ChatGPT when addressing medical questions. |
| Guo et al 2023 | Canada | Original article | Preprint | neuroGPT-X: Towards an Accountable Expert Opinion Tool for Vestibular Schwannoma | This study enhanced OpenAI's GPT-3 model for answering questions about vestibular schwannoma. The context-enriched GPT model provided faster and non-inferior responses compared to expert neurosurgeons. The model's responses were often rated higher and included in-text citations and references. However, expert surgeons expressed concerns about the model's reliability in addressing nuanced and controversial aspects of management. The study also introduced neuroGPT-X, a chat-based platform for clinical support with accurate and reliable information. |
| Gurha et al 2003 | USA | Commentary | Peer-reviewed | ChatGPT and other artificial intelligence chatbots and biomedical writing | An AI chatbot or AI writer should not be listed as an author in scientific publications because authors provide nuanced interpretation and insight beyond synthesizing sentences. Authors cannot claim ownership of text generated by chatbots and should avoid plagiarism by acknowledging their use. Chatbots are limited in generating in-depth and critical scientific writings and lack the essential insight required for review articles, commentaries, or perspectives. Large language models (LLMs) have potential in scientific writing but should be carefully evaluated for validity and edited for incorrect statements. Chatbots have various biomedical and clinical applications but require expert review and verification. LLM-based programs face challenges in extracting reliable data from biased and low-quality sources. As programs advance, commercialization may occur, but scientists should maintain access to platforms for synthesizing and extracting advanced information. JCA editors prioritize articles with nuanced insight and new understanding. |
| Hurley 2023 | USA | Commentary | Peer-reviewed | Your AI Program Will Write Your Paper Now: Neurology Editors on Managing Artificial Intelligence Submissions | ChatGPT, the language model developed by OpenAI, is creating ripples in the academic world, with several journals considering guidelines for its usage. Experts note the AI's remarkable writing ability and its potential to generate data and references, which raises concerns about authenticity. While some editors suggest there could be a place for ChatGPT to assist with language barriers and manuscript restructuring, there's consensus that the AI should not be credited as an author, and its usage should be disclosed. Additionally, experts emphasize the need for human oversight and responsibility for content generated with AI's help. |
| Haemmerli et al 2023 | Switzerland | Original article | Preprint | ChatGPT in glioma patient adjuvant therapy decision making: ready to assume the role of a doctor in the tumour | In this study, the performance of ChatGPT, an AI-based chatbot, was assessed in providing treatment recommendations for glioma patients within a multidisciplinary decision-making context. ChatGPT demonstrated accuracy in identifying cases as gliomas; however, it faced challenges when specifying tumor subtypes. The treatment recommendations and regimens generated by the tool were generally |
|  |  |  |  | board? | rated as good, while its ability to consider functional status received a moderate rating. Experts found ChatGPT's involvement in the CNS Tumor Board to be valuable, recognizing its potential for improvement and learning. |
| Harskamp and Clercq 2023 | Netherlands | Original article | Preprint | Performance of ChatGPT as an AI-assisted decision support tool in medicine: a proof-of-concept study for interpreting symptoms and management of common cardiac conditions (AMSTELHEART-2) | The AMSTELHEART-2 study evaluated the performance of the ChatGPT language model in answering cardiovascular trivia questions and interpreting case vignettes. The model achieved an overall accuracy of 74% in answering multiple-choice questions and provided appropriate advice for patients reaching out to primary care. However, it had limitations in addressing medical questions from physicians seeking expert consultation. The study's strengths include using various approaches to evaluate ChatGPT, while its limitations include a small sample size. Further refinements of the model are needed before considering its use in real-life scenarios. |
| Hill-Yardin et al 2023 | Australia | Viewpoint-Commentary | Peer-reviewed | A Chat(GPT) about the future of scientific publishing | The use of AI-generated software, such as ChatGPT, in academic publishing has sparked discussions and debates. The generated text can be accurate, logical, and grammatically correct, but it lacks a distinct "voice" and is generic. The writing style of the generated text may resemble the bland and formulaic style often found in scientific articles. Introducing more diversity in writing styles and discussing mistakes and unsupported hypotheses could help differentiate human writing from AI-generated writing. AI may eventually learn individual writing styles and inject more nuanced diction. The use of AI and predictive text software raises questions about language patterns, vocabulary, and linguistic diversity. ChatGPT's generated text is too shallow for the detailed knowledge and interpretation required in neuroscience. The lack of accountability and transparency in AI-generated text is a fundamental concern for its use in academic publishing. |
| Hirani et al 2023 | USA | Commentary | Peer-reviewed | Experimenting with ChatGPT: Concerns for Academic Medicine | The functionality of ChatGPT in academic medical publishing was examined, revealing the potential for fabrication of inaccurate research. The lack of transparency and accountability in ChatGPT's output raised concerns and prompted guidelines from publishing companies and journal editors. Bias is a significant problem as well because word embeddings, which are used in ChatGPT training, introduce biases originating from humans into the original data. Algorithmic bias in medical AI systems, which also leads to care disparities, may have a negative impact on clinical decision-making. An analysis uncovered biases in ChatGPT's responses, such as professions and jobs that are linked with particular genders. Physicians should advocate for transparency, regulation, and accountability in LLM training datasets and algorithms to address the issues of fabrication and bias. |
| Homolak 2023 | Croatia | Commentary | Peer-reviewed | Opportunities and risks of ChatGPT in medicine, science, and academic publishing: a modern Promethean dilemma | The release of ChatGPT has generated excitement about the potential of AI in various fields, including academic publishing. AI has the potential to revolutionize healthcare but is unlikely to replace physicians due to limitations in context and nuance. Ethical issues surround conversational AI in medical practice, including biased data sets and accountability for mistakes. ChatGPT's ability to generate scientific manuscripts raises concerns about reliability and the potential for misinformation. Acknowledging limitations and addressing ethical challenges are necessary before implementing AI in publishing. While AI has the potential to improve efficiency, a mindful approach and open debate about risks and benefits are essential. |
| Hosseini et al 2023 | USA | Original article | Preprint | ChatGPT versus the neurosurgical written boards: a comparative analysis of artificial intelligence/machine learning performance on neurosurgical board-style questions | A hybrid panel discussion about the use of ChatGPT in education, research, and healthcare was conducted. 420 responses were received from 844 participants, with a response rate of 49.7%. 40% of the audience had used ChatGPT, with more trainees than faculty. Interest in wider application was higher among ChatGPT users. Greatest uncertainty was around its use in education. Varied perspectives were present among different roles (trainee, faculty, staff). The need for further discussion about LLM usage was highlighted. A thoughtful, measured approach in adopting ChatGPT was suggested to reduce potential risks. |
| Howard et al 2023 | UK | Commentary | Peer reviewed | ChatGPT and antimicrobial advice: the end of the consulting infection doctor? | ChatGPT has been noted for its potential in medical scenarios, stimulating urgent discussions in the medical community. Despite its limitations, it's considered as capable as an average human physician in answering some open-ended medical queries. The study tested ChatGPT in eight hypothetical infection scenarios. It recognized natural language effectively but struggled with complex situational aspects. Its responses were coherent, clear, and reflective of its information sources. Some inconsistencies and erroneous advice emerged on repeated questioning. Deficits in situational awareness, inference, and consistency are major barriers to ChatGPT's clinical implementation. Despite no access to specific medical databases, it provides compelling responses. The authors propose a modifiable qualitative framework for future improvements. |
| Hsu et al 2023 | Taiwan | Original article | Preprint | Plagiarism, Quality, and Correctness of ChatGPT-Generated vs Human-Written Abstract for Research Paper | This study aimed to evaluate the quality and accuracy of medical abstracts generated by ChatGPT, specifically the ChatPDF version, using a selection of 20 randomised controlled trials in the field of psychiatry. Two types of abstracts, structured and unstructured, were considered.<br>The generated abstracts were analyzed in terms of similarity, plagiarism, AI-content, and subheading proportion. They were also evaluated by five experts in psychiatry for quality, using a Likert scale. Further, the validity of the conclusions<br>generated by ChatPDF was checked.<br>The findings showed a similarity (duplicate content) between the original and the generated abstracts of 16.35%. The plagiarism percentage was 18.75%, with 11.20% being self-plagiarism. After adjusting for self-plagiarism, the actual plagiarism percentage was 7.55%. The AI-content percentage was notably higher in unstructured abstracts (75.58%) compared to structured ones (31.48%).<br>The quality of generated abstracts was found to be lower than the original abstracts, especially for the unstructured format. Interestingly, a structured format and a higher H-index were associated with higher quality scores in generated and original abstracts, respectively.<br>Lastly, while the ability of experts to accurately identify original authors was fairly low (40%) for structured abstracts, it was significantly higher (73%) for unstructured ones. However, a noteworthy concern was that 30% of the abstracts generated by ChatGPT presented incorrect conclusions.<br>These findings suggest that while the performance of ChatGPT in generating medical abstracts has potential, improvements are necessary, particularly in terms of the quality of unstructured abstracts and the accuracy of conclusions. |
| Huang et al 2023 | China | Review Article | Peer-reviewed | The role of ChatGPT in scientific communication: writing better scientific review articles | Topic Selection: ChatGPT generates keywords and suggests research areas, helping scientists choose relevant topics.<br>Literature Search: The AI provides relevant search queries and database suggestions for efficient literature searches.<br>Article Selection: ChatGPT generates summaries of articles, providing context for relevance in a review.<br>Citation and Referencing: It aids in accurate citation and referencing, generating appropriate formats.<br>Efficiency: By automating tedious tasks, ChatGPT facilitates comprehensive, efficient reviews, enhancing manuscript quality.<br>Time-saving: The AI tool streamlines the writing process, saving time and effort.<br>Caution Required: Despite its benefits, scientists should exercise care, reviewing and editing generated text to avoid plagiarism and fabrication.<br>Outline Development: Scientists input topic, ChatGPT generates and organizes related subtopics into a logical outline.<br>Adding Details: ChatGPT suggests key points and relevant literature to add depth to the outline.<br>Text Improvement: Scientists input text, ChatGPT analyzes and provides suggestions for improvements in grammar, sentence structure, and style.<br>Suggested Improvements: ChatGPT offers advice to enhance clarity, precision and avoid jargon.<br>Example Provision: The AI tool provides examples of well-written scientific articles<br>or sentences to illustrate the improvements.<br>Feedback Incorporation: Scientists can apply ChatGPT's suggestions and adjust as necessary for tone and style.<br>Enhancing Quality: By adopting ChatGPT's suggestions, scientists can improve their writing's clarity, precision, and effectiveness, leading to higher quality manuscripts.<br>Assisting Non-Native English Speakers: ChatGPT aids with correct grammar, sentence structure, and appropriate vocabulary, even offering translation help.<br>Drawbacks: AI may lack context, introduce bias, encourage over-reliance, struggle with technical terms, and require substantial investment.<br>Plagiarism Risk: AI-generated text can resemble other sources; to minimize plagiarism risk, use AI as supplement, properly attribute sources, and use plagiarism-detection tools.<br>Avoiding Plagiarism: Understand text sources, use multiple references, ensure accurate citation, employ plagiarism-detection software, and carefully review and edit AI-generated text.<br>Summary: While beneficial, ChatGPT use necessitates vigilance against plagiarism and adherence to proper citation and editing practices.<br>Contextual Understanding: ChatGPT lacks contextual understanding; humans ensure generated content is appropriate for its purpose.<br>Checking Accuracy: ChatGPT can produce inaccuracies; human oversight verifies content accuracy.<br>Editing and Formatting: AI-generated text needs editing and formatting to meet article requirements; human oversight ensures compliance with these standards.<br>Conclusion: ChatGPT significantly enhances scientific writing but requires human oversight to prevent plagiarism and verify accuracy.<br>Future of AI: Advanced AI tools will further streamline scientific research processes, enabling impactful research output. |
| Janssen et al 2023 | The Netherlands | Commentary | Peer-reviewed | The use of ChatGPT and other large language models in surgical science | Surgical scientists conducting clinical trials and research on artificial intelligence in surgery are currently investigating the potential impact of Large Language Models (LLMs), such as ChatGPT, which is set to be published on November 30, 2022.<br>From the perspective of surgeon-scientists and editors, LLMs have the potential to enhance productivity and efficiency in writing tasks. They can be employed for data extraction and clinical decision-making purposes as well.<br>LLMs can assist in generating drafts for study ideas, research protocols, manuscripts, grant proposals, instructional materials, and patient education materials. They can also help improve text and rectify errors for non-native speakers.<br>Integration of LLMs into electronic health record systems could enable information extraction from patient records and other data repositories, automating data<br>collection for research purposes. This could potentially alleviate challenges associated with manual data extraction.<br>Advanced LLMs could find application in clinical workflows for evaluating patient information and generating patient management plans, leading to more efficient patient care and improved outcomes.<br>However, there are limitations and risks associated with using LLMs. These include the possibility of "neural hallucinations," where the model generates incorrect or nonsensical text, and the potential introduction of bias into the model's output.<br>Establishing precise criteria and guidelines for LLM usage is crucial to ensure their trustworthiness and security in surgical science, particularly when considering the ethical and legal implications of their use in patient management.<br>Despite these limitations, LLMs have the potential to be valuable tools in surgical science and clinical practice by augmenting human expertise rather than replacing it.<br>The article emphasizes the importance of ongoing monitoring of developments in this field and evaluating the impact of language models on surgical science. |
| Johnson et al 2023 | USA | Original article | Peer-reviewed | Using ChatGPT to evaluate cancer myths and misconceptions: artificial intelligence and cancer information | In this study, the accuracy of cancer information provided by ChatGPT, an AI chatbot, was compared to the responses from the National Cancer Institute (NCI) regarding common cancer myths and misconceptions.<br>Thirteen questions related to cancer were posed to ChatGPT, and the precision of its answers was assessed. Five scientists, who had experience in cancer treatment and false information, reviewed both the NCI's responses and ChatGPT's responses independently and blinded. The results showed that the NCI's answers received a 100% accuracy rating from all five reviewers for all 13 questions, indicating full interrater agreement. ChatGPT's answers received a 96.9% accuracy rating from all five reviewers for 11 out of the 13 questions.<br>There were minimal differences observed in word count and readability grade level between the responses from NCI and ChatGPT. The study suggests that ChatGPT provides accurate information on common cancer myths and misconceptions without disseminating misinformation or harmful information.<br>The researchers also conducted additional rounds of questioning to assess whether repetitive questioning could lead to semantic variations and potential misinformation. The answers remained consistent, indicating that the accuracy of the responses was not affected by repeated questioning. |
|  |  |  |  |  | The study underscores the positive potential of ChatGPT and AI systems in delivering accurate cancer-related information. However, it highlights the importance of monitoring and evaluating the use of such tools in online communication settings to address potential biases and health disparities.<br>Limitations of the study include the evaluation of only common cancer misinformation in English and the possibility of outdated scientific information in ChatGPT's training data.<br>Further research is needed to assess the accuracy of other chatbots and AI-driven systems, their performance in addressing diverse claims about cancer, and the development of an ideal infrastructure to monitor and ensure the accuracy of cancer information online. |
| Juhi et al 2023 | India | Original article | Peer reviewed | The Capability of ChatGPT in Predicting and Explaining Common Drug-Drug Interactions | The study aimed to investigate the effectiveness of ChatGPT in predicting and explaining common drug-drug interactions (DDIs). A total of 40 DDI pairs were used to converse with ChatGPT using a two-stage question approach.<br>The first question asked whether two specific drugs can be taken together, and the second question asked why those drugs should not be taken together. The responses from ChatGPT were checked by two pharmacologists and categorized as "correct" or "incorrect." The "correct" responses were further classified as "conclusive" or "inconclusive."<br>Among the 40 DDI pairs, one answer was incorrect in the first question, and one answer was wrong in the second question.<br>For the first question, 19 answers were classified as conclusive and 20 as inconclusive. For the second question, 17 answers were conclusive, and 22 were inconclusive.<br>Using Flesch reading ease and Flesch-Kincaid grade level, the readability ratings of the answers were assessed. The answers were found to have substantially higher grade levels than anticipated for a fictitious sixth grade, making them rather challenging to read.<br>According to the study, ChatGPT can be used to predict and explain DDIs to a limited extent. Patients without rapid access to medical facilities may receive some guidance from it, but it may also provide insufficient information. It need further work before patients might possibly use it to comprehend DDIs. |
| Kaneda 2023 | Japan | Commentary | Peer-reviewed | In the Era of Prominent AI, What Role Will Physicians Be Expected to Play? | A large language model (LLM) called ChatGPT, created by OpenAI, has become well-known for producing conversational responses.<br>An AI chat application called Chat Doctor has proven to be quite accurate at prescribing prescriptions based on talks between patients and doctors.<br>The ability of LLM models to aid patients and medical personnel in the sphere of healthcare is underlined.<br>As AI models have limits in offering the most recent research, ethical judgement,<br>and clinical intuition, doctors will continue to play an important role in healthcare settings. Collaboration between medical professionals, administrators, and stakeholders is crucial to advance healthcare reform and effectively utilize AI. |
| Kim 2022 | Republic of Korea | Original | Preprint | Search for Medical Information and Treatment Options for Musculoskeletal Disorders through an Artificial Intelligence Chatbot: Focusing on Shoulder Impingement Syndrome | The study evaluated the ability of ChatGPT, an artificial intelligence chatbot, to provide medical information and treatment options for subacromial impingement syndrome (SIS).<br>ChatGPT was able to generate responses that included correct definitions, prevalence, and risk factors of SIS based on input messages.<br>It also provided information on symptoms, diseases with similar symptoms, and orthopedic tests related to SIS.<br>ChatGPT suggested treatment options such as rest, medication, physical therapy, and potential surgical intervention.<br>However, some of the information provided by ChatGPT was biased or inappropriate depending on the individual's condition, indicating the limitations of relying solely on AI chatbots for accurate medical information.<br>The study concluded that while ChatGPT can offer overall helpful information about SIS, caution is necessary, and it still falls short in providing accurate and individualized medical information and treatment options.<br>With further advancements in natural language processing technology, it is expected that AI models like ChatGPT can deliver more detailed and precise medical information in the future. |
| Kim 2023 | China | Original | Peer reviewed | Using ChatGPT for language editing in scientific articles | The use of AI chatbots as authors in scientific articles is an ethical issue and has raised concerns in scientific journals.<br>ChatGPT, developed by OpenAI, is a popular AI-powered chatbot available for free to anyone.<br>The study submitted questions to ChatGPT regarding the effect of streptozotocin-induced diabetes on facial bone growth in rats.<br>ChatGPT provided a detailed response with references to studies, but upon verification, all the references were found to be fake.<br>ChatGPT's performance in editing English grammar was found to be excellent.<br>Language editing services like ChatGPT can be advantageous for authors, but they should not be listed as co-authors in scientific articles. |
| Koo 2023 | China | Letter to the editor | Peer-reviewed | The importance of proper use of ChatGPT in medical writing | ChatGPT has several helpful applications in the writing process, including paraphrasing difficult sentences, translation, spell-checking, grammar correction, improving clarity and flow of text, generating outlines and abstracts, and formatting references.<br>The author of the article in Radiology found that ChatGPT could write an entire article based on inputted headings and subheadings.<br>The potential risks of using AI in the writing process were discussed, emphasizing the importance of proper and ethical use of ChatGPT.<br>ChatGPT is a versatile tool that can streamline the writing process for researchers, but it should be used responsibly to maintain ethical standards. |
| Kumar 2023 | Ireland | Original | Peer- | Analysis of ChatGPT | he study aimed to evaluate the effectiveness of ChatGPT, an AI chatbot, in the |
|  |  | article | reviewed | tool to assess the potential of its utility for academic writing in biomedical domain | context of academic writing within the field of biomedical sciences. Five random topics were inputted into ChatGPT, and the study assessed factors such as response time, content quality, and reliability. The responses generated by ChatGPT were exported to a Word file and analyzed for originality using Urkund software.<br>The findings revealed that ChatGPT had a fast response rate, producing 300-500 words of text in under 2 minutes. However, the content fell short of the expected quality and depth typically found in academic writing. Limitations observed included deficiencies in word count, referencing errors, and a lack of academic merit.<br>In conclusion, while ChatGPT has the potential to serve as a useful tool for training and improving academic writing skills, it cannot replace human intelligence. When used appropriately and under academic mentoring, it can assist in refining human capabilities. Nevertheless, it is crucial to acknowledge the limitations of ChatGPT and complement its usage with human expertise to ensure the production of high-quality academic work. |
| Lee et al 2023 | USA | Review article | Peer-reviewed | Benefits, Limits, and Risks of GPT-4 as an AI Chatbot for Medicine | The use of artificial intelligence (AI) in medicine is expanding, including applications in medical image analysis, drug interaction detection, patient risk identification, and medical note coding.<br>This article focuses on a specific type of AI called a medical AI chatbot, utilizing the GPT-4 system developed by OpenAI.<br>A chatbot consists of a general-purpose AI system and a chat interface, allowing users to enter queries in natural language and receive relevant responses.<br>Prompt engineering is essential for optimizing chatbot performance, as current systems are sensitive to the wording and form of prompts.<br>While GPT-4 excels in providing correct answers to questions with firm answers, its most interesting interactions occur when prompts have no single correct response. Chatbot errors, referred to as hallucinations, can be dangerous in medical scenarios, so it is crucial to verify the output.<br>GPT-4 can help catch mistakes, not only in its own work but also in human-generated content, providing a means of error detection and correction. GPT-4 is a general-purpose AI system developed by OpenAI, designed to have broad cognitive skills rather than being programmed for specific tasks in medicine.<br>Prompts for GPT-4 can be in the form of questions or directives to perform specific tasks, and they can be written in various human languages and include data inputs like spreadsheets, research papers, and equations.<br>Microsoft Research and OpenAI have been studying the applications of GPT-4 in healthcare and medicine, focusing on areas such as documentation, data interoperability, diagnosis, research, and education.<br>Other notable AI chatbots like LaMDA (Google) and GPT-3.5 have also been explored for medical applications, despite not being explicitly trained for healthcare purposes. |
|  |  |  |  |  | Three scenario-based examples are provided to demonstrate potential medical use of GPT-4, including medical note-taking, performance on a USMLE problem, and providing advice in a "curbside consult" situation.<br>GPT-4's responses have shown improvements over time, and it has the ability to generate medical notes, answer questions, generate orders, and provide feedback to clinicians and patients.<br>However, GPT-4 is not infallible and can make errors or hallucinations, so mechanisms for error detection and correction should be implemented in future deployments. GPT-4, when tested with questions from the USMLE, answers correctly over 90% of the time, showcasing its innate medical knowledge.<br>GPT-4 can be used for medical consultation, providing useful responses and aiding in tasks like diagnosis, research, and education.<br>It can generate medical notes, provide technical analysis, summarize research, identify prior work, and ask follow-up research questions.<br>GPT-4 has the potential to write computer programs, translate languages, decipher medical documents, and provide emotional support.<br>While GPT-4 has limitations and can make mistakes, it also has the ability to catch mistakes made by both AI and humans.<br>The evaluation of GPT-4's general intelligence and the level of trust users can place in it remain important questions for further discussion.<br>GPT-4 is a powerful tool with evolving capabilities, and its responsible use can bring new possibilities while acknowledging associated risks. |
| Levin et al 2023 | Canada | Original articles | Peer-reviewed | ChatGPT-written OBGYN abstracts fool practitioners | The goal of the study was to determine how well doctors with different degrees of publication experience could distinguish between abstracts created by ChatGPT and human-written abstracts.<br>In the study, 20 abstracts—10 authored by ChatGPT and 10 by humans—were submitted for examination by eight reviewers.<br>30% of ChatGPT-written abstracts and 43.7% of human-written abstracts were correctly identified by the reviewers. The accuracy in identifying ChatGPT-written abstracts was associated with publication experience rather than years of experience.<br>A modified Grammarly-based tool showed higher accuracy in identifying abstracts compared to the reviewers.<br>Web-based AI output detectors were able to detect ChatGPT text, suggesting their potential use in screening submissions to journals.<br>The study highlights the need for improved means of detecting ChatGPT-written scientific abstracts.<br>The limitations of the study include a small sample size of reviewers and referencing a preprint report, while the novelty and total number of abstracts tested are considered strengths.<br>Efforts should be focused on optimizing methods for the detection of ChatGPT-written abstracts. |
| Li et al 2023 | USA | Commentary | Peer- | Ethics of large language | The medical industry has become interested in large language models (LLMs), |
|  |  |  | reviewed | models in medicine and medical research | such as ChatGPT, because they may be utilised for text production, summarization, and correction activities.<br>LLMs can produce content for patient-friendly communication, standardised reporting, presentation outlines, sample cover letters, and clinical documentation. As LLMs mirror the training data, which may perpetuate prejudices and underrepresent some perspectives, ethical problems include the possibility of hidden biases in LLMs.<br>Trust and credibility of LLM-generated content are challenges, as LLM outputs are difficult to trace and may be inaccurate. Current models do not evaluate quality or provide measures of uncertainty.<br>Authorship becomes a complex issue when LLMs are used, as LLMs lack intentionality and cannot approve or be held accountable for the final work. New guidelines may be required.<br>Varying payment models for LLMs may widen digital divides, and affordability and accessibility should be ensured.<br>Privacy concerns arise regarding the collection, use, and potential dissemination of data inputted into LLMs. Strict controls and informed consent are necessary.<br>It is important to establish guidelines that promote responsible and effective use of LLMs, rather than imposing an outright ban on their use.<br>In summary, while LLMs have the potential to revolutionize medicine and medical research, careful consideration of their biases, trustworthiness, authorship, equitability, and privacy implications is crucial. Responsible guidelines should be established to ensure their ethical use. |
| Loh 2023 | Australia | Review article | Peer-reviewed | ChatGPT and generative AI chatbots: challenges and opportunities for science, medicine and medical leaders | ChatGPT, an AI chatbot tool released by OpenAI, has gained significant popularity and raised concerns about fraud and plagiarism due to its ability to generate text similar to human-authored content.<br>The risks of generative AI tools in health include challenges of fraud and plagiarism in educational settings, the potential for biased outputs, and privacy and cybersecurity concerns.<br>AI tools like ChatGPT have been used to generate scientific articles, leading to changes in guidelines by scientific meetings and publishers to address authorship and credibility.<br>Despite the risks, generative AI tools offer opportunities for scientific research, including assistance in experimental design, peer review, and administrative tasks such as grant applications and research article editing.<br>In clinical practice, AI chatbots like ChatGPT can support frontline clinicians by automating administrative tasks, such as generating discharge summaries and medical letters.<br>AI chatbots can also assist in patient communication by providing accurate information and answering low-risk health questions, although ethical considerations need to be addressed.<br>Some health systems have already started using generative AI technology to improve healthcare provision and alleviate administrative burdens and clinician<br>burnout. |
| Li 2023 | China | Commentary | Peer reviewed | ChatGPT has made the field of surgery full of opportunities and challenges | ChatGPT is a language generation model developed by OpenAI and is capable of generating high-quality text content.<br>It has been trained on a large amount of preprocessed and filtered text data.<br>IBM's Watson robot is an AI technology specifically designed for the medical field, providing diagnosis and treatment suggestions based on clinical guidelines.<br>In the field of surgery, the Da Vinci robotic surgical system is widely used, but surgeons still operate the surgeries and AI technology assists in surgical planning, navigation, and support.<br>While ChatGPT can provide valuable diagnostic and treatment suggestions, doctors must evaluate and judge these suggestions and comply with legal regulations.<br>ChatGPT is suitable for document polishing and has the potential to be a powerful medical assistant in the future.<br>The current application of ChatGPT in the medical field may not have a disruptive effect yet, but its potential is promising and it can become a valuable tool with further development and training. |
| Liebrenz et al 2023 | UK | Commentary | Peer reviewed | Generating scholarly content with ChatGPT: ethical challenges for medical publishing | The use of generative AI, such as ChatGPT, in medical publishing practices has unknown implications, but it raises substantial ethical concerns.<br>ChatGPT is an AI chatbot developed by OpenAI that generates responses based on internet sources and has been used to create university essays and scholarly articles.<br>The imperceptibility of AI-generated content to human readers and anti-plagiarism software raises concerns about copyright, attribution, plagiarism, and authorship.<br>There is a debate about whether AI can fulfill the criteria for authorship set by organizations like the International Committee of Medical Journal Editors.<br>Organisations like the Committee on Publication Ethics and the International Association of Scientific, Technical, and Medical Publishers have given ethical principles and guidelines for AI in publishing.<br>As AI technologies proliferate, comprehensive talks about authorship laws and AI-generated work are seen as urgent and crucial.<br>The usage of ChatGPT and comparable AI platforms might result in a paywall-based business model, escalating already-existing inequities in scientific publication. ChatGPT's accessibility and ease of use have the potential to increase scholarly output and democratize knowledge dissemination in multiple languages.<br>However, there are concerns about the potential harm caused by ChatGPT in producing misleading or inaccurate content, contributing to scholarly misinformation.<br>Competitors to ChatGPT may arise, amplifying the challenges associated with AI-generated content in scholarly publishing.<br>The Lancet Digital Health and the Lancet family are called upon to carefully consider the ethical implications of publishing articles produced by AI and initiate discussions on comprehensive guidance for AI-generated content in scholarly<br>publishing. |
| Lin 2023 | Hong Kong | Commentary | Preprint | Modernizing authorship criteria: Challenges from exponential authorship inflation and generative artificial intelligence | The conventional requirement for authorship, based on "substantial contributions," is out of sync with current practices and emerging trends in scientific research. The exponential increase in the number of authors per paper poses challenges to the notion of "substantial contributions," as each author's contribution becomes increasingly small.<br>The rise of generative AI tools, like ChatGPT, in scientific research raises questions about whether they should be listed as co-authors when they substantially contribute to a paper.<br>The International Committee of Medical Journal Editors (ICMJE) and other authoritative guidelines emphasize substantial contributions to the research and writing process as criteria for authorship.<br>However, the current criteria do not align with the increasing number of authors and the potential substantial contributions of AI tools.<br>A new approach is proposed, where authorship is seen as proportional, role-specific credit, reflecting the specific contributions made by individuals in various roles.<br>The CRediT system, which defines specific roles in research, can be used to specify and quantify individual contributions in a standardized manner.<br>Organizations, publishers, and authors should consider reevaluating and updating authorship criteria to better reflect current research practices and the involvement of generative AI tools.<br>Recommendations include revising the requirement of "substantial contributions," requiring authors to specify contributions using a standardized system like CRediT, and actively documenting and discussing contributions throughout the research process. |
| Maeker and Maeker-Poquet 2023 | France | Commentary | Peer-reviewed | ChatGPT: A solution for producing medical literature reviews? | Artificial intelligence (AI) can produce grammatically correct and coherent text based on patterns it has learned, but it lacks a deep understanding of the content.<br>AI like ChatGPT operates by processing natural language and following patterns in data, not by truly understanding the content.<br>AI's capability to synthesize complex thoughts or scientific content is limited due to its lack of true understanding.<br>An experiment was conducted to see if ChatGPT could write a literature review, drawing upon its learning from diverse sources including PubMed.<br>The initial trial showed that while ChatGPT could provide clear, well-written content, it lacked sufficient references, often invented content, and did not meet established literature review guidelines (e.g., PRISMA and ICMJE).<br>A second trial, with more structured input and guidance, led to more coherent<br>content but still had shortcomings in terms of depth, reference handling, and staying on topic.<br>Despite the limitations, ChatGPT could be a useful tool in drafting early versions of literature reviews or helping novice writers get started, owing to its speed and ability to generate content.<br>AI could also help suggest initial manuscript structure, rephrase and summarize content, and suggest references, although it can only draw from information up to 2021 and can't compare and interpret results.<br>There are ethical considerations, such as plagiarism detection and authorship issues, that arise when using AI tools for writing scientific literature.<br>Guidelines set by publishers like Elsevier allow for the use of AI for improving readability and language but discourage it from replacing key research steps like producing scientific information, analyzing data, or formulating conclusions.<br>The use of AI such as ChatGPT in writing scientific literature has potential, but should be considered a supplement to, not a replacement for, human expertise and judgement. Ethical issues around this usage remain under debate. |
| Marchandot et al 2023 | France | Commentary | Peer-reviewed | ChatGPT: the next frontier in academic writing for cardiologists or a pandora's box of ethical dilemmas | ChatGPT, developed by OpenAI, is a powerful language model capable of generating human-like text. It can understand context and generate coherent, fluent text that is often indistinguishable from text written by humans.<br>The model has potential applications in academic research, such as assisting in data analysis, literature reviews, and drafting research papers. It can analyze vast amounts of data quickly and can be trained on specific topics to help find and summarize relevant literature.<br>ChatGPT can assist in revising scientific manuscripts, identifying and correcting grammar and spelling errors, suggesting alternative phrasing, and even proposing additional experiments or data analysis.<br>There are potential downsides to using ChatGPT in academic research. For example, if trained on biased data, the model might produce biased results. It may also not understand the nuances of a specific field and produce inaccurate results.<br>Dependence on models like ChatGPT could lead to a decrease in critical thinking and creativity among researchers. There are also ethical concerns related to plagiarism, as the model may generate text similar to existing content.<br>There is ongoing debate on the use of AI tools like ChatGPT in academic research. |
|  |  |  |  |  | It's controversial whether it's appropriate to cite ChatGPT as an author in published literature.<br>Despite concerns and potential downsides, the authors of the commentary acknowledge the invaluable assistance of ChatGPT in writing and editing the text, advocating for its recognition as a co-author. They call for academic institutions and publishers to establish guidelines for using AI-generated text in academic research. |
| Martínez-Sellés and Marina-Breysse 2023 | Spain | Review article | Peer-reviewed | Current and Future Use of Artificial Intelligence in Electrocardiography | AI technology is rapidly being integrated into electrocardiography (ECG) for the detection, categorization, and treatment of heart disease.<br>AI algorithms have the ability to assist clinical professionals in analyzing and identifying ECG abnormalities such as arrhythmias, ST-segment changes, and QT prolongation.<br>AI can be utilized alone or in combination with clinical factors to predict the occurrence of arrhythmias, sudden cardiac death, stroke, and other cardiovascular events.<br>Real-time monitoring of ECG signals from wearable electronic devices and cardiac implants can be facilitated by AI, alerting medical professionals or patients when significant changes occur based on timing, duration, and context.<br>By reducing noise, artifacts, and interference, as well as extracting data that may not be visible to the human eye, such as heart rate variability, beat-to-beat intervals, and wavelet transforms, AI can enhance signal processing, leading to improved ECG quality and accuracy.<br>AI can support therapy decision-making by assisting with patient selection, treatment optimization, timing of symptom-to-treatment, and cost-effectiveness. Furthermore, by predicting the outcome of antiarrhythmic drug or cardiac implanted device therapy, AI can reduce the risk of cardiac toxicity and facilitate the early activation of emergency protocols in patients with ST-segment elevation.<br>The fusion of ECG data with information from other modalities, including imaging, genomics, proteomics, and biomarkers, can be facilitated by AI.<br>As more data becomes available and AI algorithms continue to advance, it is expected that AI will play an increasingly significant role in the diagnosis and management of ECG-related conditions in the future. |
| Mehnen et al 2023 | Austria | Original article | Preprint | ChatGPT as a medical doctor? A diagnostic accuracy study on | The study tested the diagnostic accuracy of ChatGPT using 50 clinical case vignettes, including 40 common complaints and 10 rare diseases. |
|  |  |  |  | common and rare diseases | The case vignettes were collected from previous research and from the first matching articles on PubMed for randomly selected rare disorders with an approved orphan drug.<br>ChatGPT was prompted with the phrase “What are the 10 most likely diagnoses for this patient?” followed by the full text of the clinical case vignette.<br>Each vignette was prompted three times in independent chats using both ChatGPT 3.5 and ChatGPT 4, totaling 300 prompts and 3000 suggested diagnoses.<br>The 50 case vignettes were also presented to three human medical doctors for diagnosis without the use of search engines.<br>The correctness of suggested diagnoses was assessed by comparing them with the correct diagnoses provided in the case vignettes.<br>For common cases, ChatGPT 3.5 solved more than 90% of all cases within the top two suggested diagnoses, while ChatGPT 4 solved 100% of all cases within the top three suggested diagnoses.<br>For rare cases, ChatGPT 3.5 achieved an accuracy of 60% within the top 10 suggestions, while ChatGPT 4 achieved an accuracy of 90% within eight or more suggestions.<br>Fleiss' kappa was used for diagnosis agreement hypothesis-testing between different versions of ChatGPT and the correct diagnoses, indicating a significant similarity to the correct diagnoses.<br>Despite the high diagnostic accuracy on common cases, caution should be exercised when using ChatGPT for medical advice, especially for non-professionals, due to the lower performance on rare cases. |
| Mello and Guha 2023 | USA | Commentary | Peer-reviewed | ChatGPT and Physicians' Malpractice Risk | Large language models (LLMs) like ChatGPT have the potential to augment or replace human decision-making in various fields, including medical decision-making. However, they also bring with them a risk of liability for practitioners who use them.<br>The legal position of physicians using these models is unclear. However, the precedent of clinical practice guidelines may provide some guidance. In cases where physicians have followed these guidelines, it has often constituted exculpatory evidence in malpractice suits.<br>LLMs have some distinctive features that make them different from clinical practice guidelines. They have a tendency to generate incorrect outputs (hallucinations),<br>often without clear references for physicians to assess the reliability of the information.<br>Most LLMs are trained on a broad array of web text, which does not discern between reputable medical articles and less reliable sources such as online forums. Efforts are being made to train LLMs on authoritative medical texts, but these are still in their infancy.<br>LLMs' outputs are not fixed, they can yield different responses for the same query, and also vary depending on the date and the phrasing of the question. This can make it difficult to determine the reasonableness of a physician's actions based on the LLM's output.<br>While clinical decision support tools often undergo rigorous validation processes, the same can't always be said for LLMs. They are mainly validated as text generators and their design and evaluation is often performed by computer scientists, not clinicians.<br>Despite these drawbacks, LLMs have their benefits. They can provide more patient-specific recommendations than other decision-support tools and may also suggest diagnostic and treatment possibilities that may be overlooked by physicians.<br>There are studies evaluating LLMs' performance in clinical scenarios, and although the results show some accuracy, there are also instances where they provide completely incorrect answers.<br>At the present time, physicians should only use LLMs to supplement more traditional forms of information seeking, such as searching for reputable sources on Google or using clinical decision support systems.<br>Future reliable LLMs may likely come from specialized systems rather than generalist systems like ChatGPT. These specialized systems can be validated more easily and are less prone to the problems of indiscriminate information sourcing.<br>While the potential of LLMs is promising, it is important to carefully understand and manage the risks associated with their use in medical practice. Continued research and development is needed to improve these systems for practical clinical use. |
| Mese 2023 | Turkey | Letter to the editor | Peer reviewed | The imperative of a radiology AI deployment registry and the potential of ChatGPT | To promote safety, efficient use, and ongoing development, the study in Clinical Radiology suggests setting up a UK registry for radiology artificial intelligence (AI) applications. |
|  |  |  |  |  | The writers of the article propose a precise description of the registry's objective, target market, and domain, as well as frequent evaluations for preserving its worth and applicability.<br>Due to its remarkable natural language interpretation and generation capabilities, ChatGPT, developed by OpenAI, can be a useful tool in this situation.<br>By condensing voluminous material and highlighting salient aspects, ChatGPT can promote more effective talks and decision-making processes between stakeholders.<br>In order to raise awareness of the register among members of the Royal College of Radiologists (RCR) and other interested parties, ChatGPT can also provide precise, coherent, and correct text.<br>By evaluating textual information like as user comments, performance reports, and research articles, ChatGPT can be helpful in the continuing monitoring and assessment of AI applications in the registry. This information can help pinpoint possible areas for improvement or concern.<br>The AI model's comprehension of English may also help with the registry's long-term upkeep by detecting pertinent updates, summarising fresh research, and creating rules to make sure the registry is kept current.<br>The incorporation of ChatGPT in the creation and upkeep of the registry may simplify monitoring, assessment, and communication procedures, potentially enhancing patient safety, resource effectiveness, and learning retention.<br>The introduction of ChatGPT into the radiology AI deployment registry has the potential to make a substantial impact on the development of AI technology. |
| Nastasi et al 2023 | USA | Commentary | Preprint | Does ChatGPT Provide Appropriate and Equitable Medical Advice?: A Vignette-Based, Clinical Evaluation Across Care Contexts | A huge language model from OpenAI called ChatGPT has demonstrated amazing ability in conversational activities like successfully choosing the right radiological studies, writing research papers, and passing medical licencing examinations.<br>However, due to a lack of publicly available training data and unsolved concerns regarding the safety, fairness, and regulation of AI systems, there are doubts about its capacity to offer competent and equitable medical advice in a variety of clinical scenarios.<br>The study evaluated the clinical appropriateness of ChatGPT's answers to questions about seeking guidance across a range of settings and demographics.<br>It was discovered that ChatGPT alters its responses depending on the patient's<br>insurance status, race, and gender, frequently resulting in varied and clinically unsuitable recommendations that, if followed, could worsen health inequities.<br>The responses lacked the personalised nuance or follow-up questions that a clinician would generally offer, even though the AI frequently gave useful recommendations. Concerns concerning reliability were raised when it was discovered that responses varied greatly and were inconsistent.<br>The study's limitations include the fact that just three distinct clinical settings were tested, and that within-vignette variance was not measured.<br>Despite these drawbacks, the study offers crucial data that contextualises ChatGPT's capacity to deliver accurate and impartial guidance along the treatment continuum.<br>Conclusion: While ChatGPT can offer generic information on clinical subjects, it is currently unable to provide reliable personalised or suitable medical advice. Future improvements and training might help future LLMs give better medical recommendations. |
| Nguyen and Costedoat-Chalumeau 2023 | France | Commentary | Peer-reviewed | Artificial intelligence and internal medicine: The example of hydroxychloroquine according to ChatGPT | Artificial intelligence (AI), specifically deep learning, is revolutionizing various fields, including medicine.<br>ChatGPT is a conversational AI system developed by OpenAI, capable of providing natural and fluent communication with users.<br>ChatGPT has been pretrained on a vast amount of textual data, enabling it to generate precise and relevant responses.<br>ChatGPT can be used as an additional source of information for patients seeking medical information on topics like hydroxychloroquine (HCQ).<br>While ChatGPT's responses can be informative, they should be validated and supplemented with information from reliable sources.<br>Patients increasingly rely on the internet for medical information, and conversational AI systems like ChatGPT can provide insights into the content patients may encounter online.<br>The example of HCQ information provided by ChatGPT demonstrates both accurate and inaccurate information, highlighting the need for caution and verification.<br>ChatGPT can also be used by prescribers to obtain information on topics like dosing and monitoring requirements.<br>However, prescribers should consult healthcare professionals for precise dosing and specific guidelines.<br>Conversational AI systems have the potential to improve patient education and enhance healthcare delivery, but their limitations and potential risks should be considered.<br>Further research and development are needed to refine and enhance the<br>capabilities of conversational AI in the field of medicine. |
| Nógrádi et al 2023 | Hungary | Original article | Preprint | ChatGPT M.D.: Is there any room for generative AI in neurology and other medical areas? | This preprint research paper discusses the use of ChatGPT, an AI language model, for diagnosing neurological disorders. The study involved 12 medical doctors from the University of Szeged, Hungary, who were divided into two groups: neurologist specialists and general medical doctors. Synthetic cases representing various neurological diseases were generated by neurological experts and fed into ChatGPT to obtain diagnostic predictions.<br>The results showed that ChatGPT's diagnostic accuracy was higher than that of the general medical doctors but lower than that of the neurologist specialists. However, when considering the three most likely diagnoses or the five most likely diagnoses provided by ChatGPT, the accuracy reached a level comparable to the experts. The study also found that ChatGPT performed equally well in diagnosing acute neurological disorders compared to human doctors, but its accuracy decreased for non-acute cases.<br>The researchers observed a significant overlap in incorrect diagnoses between ChatGPT and human doctors, indicating similarities in diagnostic errors. In a subset of unsolved cases where even the experts failed to provide an accurate diagnosis, ChatGPT's most probable diagnosis matched the original diagnosis in 40% of cases, and the five most probable diagnoses included the correct diagnosis in 60% of cases.<br>The paper emphasizes that ChatGPT should be used as an augmentation tool in healthcare, with all suggestions made by the AI further evaluated by medical experts. It also highlights the potential of AI, such as ChatGPT, in supporting the triaging process for acute neurological scenarios and diagnosing rare and atypical cases.<br>The study acknowledges the ethical, integrity, and data safety concerns associated with using AI in medicine. While AI has the potential to improve healthcare, careful consideration of these concerns is necessary. |
| North 2023 | UK | Commentary | Peer reviewed | Plagiarism re-imagined | The author of this personal account reflects on a time when scientific writing was done manually, with quills, pens, and typewriters. Manuscripts were often typed by someone else, and research was conducted using limited resources like tape recorders and index cards. The author received a request to review a manuscript from the journal Life Sciences, which turned out to have significant portions copied verbatim from a paper the author had previously published. The author contacted the editor, who took the matter seriously and barred the authors from submitting further work to the journal.<br>Later, the author came across another paper published in Neuroscience Letters, which also contained substantial portions copied from the author's earlier work. The<br>author contacted the editor, who published a disclaimer acknowledging the overlap and the prior publication of the author's work. The author reflects on the irony of receiving a plagiarized manuscript for review and the increased ease of plagiarism in the modern age with AI-based text generators like ChatGPT. The author also highlights the detrimental impact of simplistic numerical methods, such as the h-index, on scientific integrity and the motivation behind academic dishonesty. |
| Oh et al 2023 | Korea | Original article | Peer reviewed | ChatGPT goes to the operating room: evaluating GPT-4 performance and its potential in surgical education and training in the era of large language models | The goal of this study was to assess how well the ChatGPT language model performed using a dataset of questions from the Korean general surgery board exam. 280 questions from the exam's first stage between 2020 and 2022 made up the dataset. The dataset did not include any questions that requested visual data. For model testing, the study used ChatGPT versions GPT-3.5 and GPT-4.<br>The study findings revealed a notable disparity in performance between the GPT-3.5 and GPT-4 models, with GPT-4 exhibiting significant improvement. GPT-4 achieved an overall accuracy rate of 76.4%, surpassing the accuracy rate of GPT-3.5, which was 46.8%. The accuracy rates varied across different subspecialties, with transplantation, breast, and hepatobiliary and pancreas demonstrating higher accuracy rates.<br>The study highlights the potential of language models like ChatGPT in comprehending complex surgical clinical information. It emphasizes the significance of the findings, as the models achieved high accuracy without fine-tuning and using prompts exclusively in the Korean language.<br>These results suggest that language models can have implications for surgical education and training, providing a tool for evaluating surgical knowledge and skills. However, further research and fine-tuning of the models are needed to enhance their performance and applicability in the surgical field. |
| Okan 2023 | Turkey | Commentary | Peer reviewed | AI and Psychiatry: The ChatGPT Perspective | Modern language model ChatGPT, created by OpenAI, simulates human conversation and offers precise information and responses on a range of subjects.<br>A clinical decision support tool, medical content generator, drug information retrieval tool, medical translator, and patient educator are just a few of the potential uses for ChatGPT in the healthcare industry.<br>ChatGPT can support the selection of treatments and long-term patient monitoring in the field of psychiatry. It can also help with the diagnostic process, support psychiatric education, simulate case examples, assist researchers in data collecting and analysis, and more.<br>The diagnosis, treatment, and monitoring of psychiatric diseases could be revolutionised by integrating artificial intelligence, natural language processing, and GPT technologies. |
|  |  |  |  |  | These technologies can help psychiatrists make more accurate diagnoses, design effective personalized treatments, improve clinical trial accuracy and speed, and increase accessibility to mental health care through chatbots and virtual assistants. The use of AI in psychiatry has the potential to advance the field and improve patient outcomes.<br>AI technologies like ChatGPT have exceeded expectations and are expected to provide significant support under human control. The future of psychiatry and mental health care looks promising with the continued evolution and improvement of AI technologies in the field. |
| Parsa and Ebrahimzadeh 2023 | USA | Editorial | Peer reviewed | ChatGPT in Medicine; a Disruptive Innovation or Just One Step Forward? | ChatGPT, developed by OpenAI, has gained immense popularity as a fluent chatbot with over 100 million users. Incorporating AI technologies like ChatGPT in medical education has great potential, improving patient education, health literacy, and facilitating clinical workflows.<br>ChatGPT can assist with patient inquiries, help with writing medical notes and discharge summaries, aid in decision-making and treatment planning, and support medical educators in various tasks. It can generate creative ideas, write essays and homework assignments, and even contribute as an author on research papers.<br>Challenges exist with the use of ChatGPT, such as the need for access to current information, potential biases in the training data, privacy concerns, and issues related to authorship in academic papers. Regulations and control mechanisms should be established to address ethical utilization of ChatGPT and similar tools in medicine. Despite challenges, AI tools like ChatGPT are expected to be widely adopted in the medical field in the future. |
| Patel et al 2023 | USA | Editorial | Peer reviewed | ChatGPT: friend or foe | ChatGPT, developed by OpenAI, has garnered significant attention and excitement due to its ability to generate text in response to prompts. It has demonstrated potential applications in healthcare, such as generating patient discharge summaries and simplifying radiology reports, but errors and limitations have been observed.<br>OpenAI acknowledges that ChatGPT's output can be incorrect, biased, and respond to harmful instructions, despite implementing guardrails to mitigate risks. Concerns have been raised regarding algorithmic bias, the spread of misinformation, and the potential for introducing errors or plagiarized content into scientific publications.<br>The World Association of Medical Editors has issued recommendations on the use of ChatGPT and other chatbots in scholarly publications, emphasizing the need for new tools to detect AI-generated content. Editorial policies are evolving to address the use of AI in scientific writing, including the requirement for manual checks of AI-generated output and the exclusion of AI<br>tools as authors or co-authors.<br>Careful and responsible use of ChatGPT, along with investment in robust AI output detectors, is necessary to prevent potential negative impacts on society and scholarly publishing.<br>While ChatGPT is considered a game changer, there is a need for more forethought, oversight, and technological advancements before its widespread adoption. |
| Pourhoseingholi et al 2023 | USA | Editorial | Peer reviewed | Does chatGPT (or any other artificial intelligence language tool) deserve to be included in authorship list? | ChatGPT, developed by OpenAI, is a chatbot powered by an AI neural network that can provide responses to user prompts.<br>It has been used in various fields, including healthcare, where it can assist with tasks such as accessing medical records and providing patient services.<br>ChatGPT has also been shown to be a useful tool for education and research, aiding in learning and generating high-quality manuscripts.<br>The question of whether ChatGPT should be considered an author in scientific publications has sparked debate and ethical concerns.<br>ChatGPT is seen as a tool rather than a collaborator, and it lacks the ability to participate in the research process and take responsibility for the work.<br>International guidelines for authorship criteria, such as those from ICMJE and COPE, are not met by ChatGPT.<br>The placement of ChatGPT as an author, the issue of consent, and conflicts of interest need to be addressed.<br>Policies should be implemented to prevent the inclusion of ChatGPT as an author, and transparent guidelines should be established by publishers and journals.<br>International organizations like ICMJE and COPE should consider adapting their criteria to address AI authorship.<br>Prohibition of the use of ChatGPT in research seems impractical, and efforts should focus on developing clear policies and criteria for AI authorship. |
| Rao 2023 | India | Letter to editor | Peer-reviewed | The Urgent Need for Healthcare Workforce Upskilling and Ethical Considerations in the Era of AI-Assisted Medicine | Effective implementation of AI in healthcare necessitates substantial training and upskilling of healthcare providers to ensure their proficiency in utilizing these technologies.<br>Addressing ethical and privacy concerns associated with AI in healthcare requires the establishment of stringent guidelines and robust safeguards to protect patient data.<br>Language generation models such as ChatGPT have the capability to assist healthcare professionals in composing discharge summaries, leading to improved efficiency and accuracy in documentation.<br>The responsible and conscientious utilization of AI technologies can enhance healthcare outcomes and alleviate the workload burden on healthcare providers who often face significant demands. |
|  |  |  |  |  | The integration of AI in India's healthcare system holds the potential to bring about transformative changes in healthcare delivery. However, it is crucial to exercise caution and consider all relevant factors to ensure a well-planned and regulated implementation process. |
| Ray and Majumder 2023 | India | Letter to editor | Peer-reviewed | AI Tackles Pandemics: ChatGPT's Game-Changing Impact on Infectious Disease Control | The research examines ChatGPT's possible uses as an AI language model for infectious disease management and control. ChatGPT should be regarded as an additional tool for clinical practise rather than a replacement for healthcare providers' knowledge. The problems of ChatGPT's accessibility, lack of human interaction, and inaccurate information are all acknowledged.<br>ChatGPT can assist in disseminating accurate and up-to-date information about infectious diseases, monitoring outbreaks, and aiding in diagnosis, treatment, and research.<br>Limitations of ChatGPT include potential inaccuracies from biased or incorrect input data, lack of human interaction, ethical considerations, and language barriers. Optimization of ChatGPT can be achieved through continuous data collection, refining training data, and validation by human experts.<br>Making ChatGPT more accessible can be achieved by expanding language capabilities and incorporating translation features.<br>Ethical guidelines and regulations are needed to ensure responsible use of ChatGPT, fostering accountability, providing transparency, and protecting privacy. The effectiveness of ChatGPT in managing infectious diseases can be increased through seamless interaction with current healthcare infrastructure and operations. The potential for ChatGPT and other models to play a substantial role in healthcare is made possible by the ongoing advancements in AI technology, but strict oversight and regulation are required. |
| Ros-Arlanzón and Pérez-Sempere 2023 | Spain | Letter to editor | Peer-reviewed | [ChatGPT: a novel tool for writing scientific articles, but not an author (for the time being)] | ChatGPT, developed by OpenAI, is a language model that has sparked significant debate due to its potential applications in various fields, including scientific writing and medicine.<br>ChatGPT can generate coherent and relevant text, making it a valuable tool for researchers in neurology and other disciplines.<br>However, it is important to note that ChatGPT cannot be considered an author of a scientific article. Authorship is reserved for individuals who have made substantial contributions to the research, and as an AI language model, ChatGPT cannot contribute to the conception, design, execution, analysis, or interpretation of the research.<br>ChatGPT can be acknowledged in the acknowledgments section of a scientific article or recognized as a tool used in the research process, but it should not replace human contribution in scientific research and writing.<br>The continuous development of AI technology may lead to future considerations regarding the role of ChatGPT and other AI models as coauthors or even as the main author of scientific articles. |
|  |  |  |  |  | The potential of AI, including ChatGPT, is significant, but it is important to recognize the current limitations and the need for researchers to conduct the research and take responsibility for the content of scientific publications. |
| Sabry Abdel-Messih and Kamel Boulos 2023 | Portugal | Letter to editor | Peer-reviewed | ChatGPT in Clinical Toxicology | Since its public launch on November 30, 2022, ChatGPT, developed by OpenAI in San Francisco, has made significant strides in the medical field, despite not being explicitly trained in medicine.<br>Powered by deep learning artificial intelligence (AI) techniques and based on the GPT-3 architecture, ChatGPT provides human-like responses to natural language queries.<br>Integration of ChatGPT into the Microsoft Bing search engine is currently underway, which will make it accessible to a wide range of online users worldwide, including clinicians, medical and nursing students, as well as patients.<br>The authors of a study conducted a test on ChatGPT using a clinical toxicology vignette focused on acute organophosphate poisoning, a relatively straightforward case that professionals in the field are unlikely to overlook.<br>ChatGPT successfully answered all questions related to the case, offering well-explained reasoning. Both the initial response and a regenerated response provided satisfactory results.<br>While ChatGPT demonstrated competence with straightforward cases, the authors highlight that the true challenge in clinical medicine often lies in accurate history taking, identification of signs, and eliciting pertinent information—areas where junior clinicians may encounter difficulties.<br>There is potential for future iterations of ChatGPT, specifically adapted for medical use, to be valuable in handling less common clinical cases that experts may sometimes overlook.<br>The authors envision a future where AI serves as a tool utilized by clinicians, rather than a replacement for them. They predict that in the coming years, clinicians who incorporate AI into their practice will replace those who do not. |
| Sallam 2023 | Jordan | Review | Preprint | The Utility of ChatGPT as an Example of Large Language Models in Healthcare Education, Research and Practice: Systematic Review on the Future Perspectives | A thorough analysis was done to determine the possible benefits and drawbacks of the "ChatGPT" AI-based large language model (LLM) in healthcare instruction, study, and practise.<br>The review employed PRIMSA standards and utilised the term "ChatGPT" to search English records in PubMed/MEDLINE and Google Scholar. Any preprints or published studies considering ChatGPT in the context of healthcare were eligible |
|  |  |  |  | and Potential Limitations | for inclusion.<br>60 records out of a total of 280 were deemed eligible for inclusion after a careful screening process.<br>ChatGPT was mentioned in 85% of the records that were collected, with its use in scientific writing and medical research being the most common. It was praised for its ability to reduce costs, facilitate paperwork, enable personalised therapy, and promote more health literacy in healthcare practise.<br>Nearly 97% of the records mentioned worries or potential hazards related to ChatGPT. The most frequent problems were ethical ones, including bias risk, plagiarism, copyright issues, transparency problems, legal concerns, a lack of originality, erroneous responses, a lack of understanding, and improper citations.<br>Despite these reservations, ChatGPT demonstrates promising applications that have the potential to revolutionise healthcare research, teaching, and practise. These apps should be used cautiously, though.<br>ChatGPT could be a useful resource for individualised learning in the field of health education and promote a move towards critical thinking and problem-based learning.<br>ChatGPT could improve personalised medication and streamline workflows in the healthcare industry.<br>ChatGPT could speed up the experimental design process in scientific research and improve the equity and adaptability of the research.<br>ChatGPT does not meet the criteria to be an author in scientific journals as of right now. However, modifications to the ICMJE/COPE standards may cause this to change.<br>The study urges the creation of a code of ethics and conduct for the responsible use of ChatGPT and other LLMs, engaging all parties involved in healthcare education, research, and practise. |
| Sallam et al 2023 | Jordan | Review | Peer reviewed | ChatGPT Output Regarding Compulsory Vaccination and COVID-19 Vaccine Conspiracy: A Descriptive Study at the Outset of a Paradigm | ChatGPT could be a useful resource for individualised learning in the field of health education and promote a move towards critical thinking and problem-based learning.<br>In light of its impending integration into web search engines, the study sought to assess the output of the AI-based language model ChatGPT in relation to COVID-19 vaccine conspiracy theories and mandatory vaccination. |
|  |  |  |  | Shift in Online Search for Information | On January 14, 2023, the investigation was carried out utilising ChatGPT, an OpenAI creation. Two writers assessed the correctness, clarity, conciseness, and bias of the ChatGPT-generated responses.<br>Findings revealed that ChatGPT replies rejected conspiracy theories regarding the origins of the SARS-CoV-2 virus as untrue and devoid of scientific support. Additionally, ChatGPT answers substantially refuted the claims of COVID-19 vaccine conspiracies.<br>ChatGPT exhibited objectivity when discussing mandatory vaccination, presenting both the benefits and drawbacks of such a policy. The benefits included preserving herd immunity, preventing the spread of disease, being cost-effective, and having a legal obligation. The drawbacks included moral and legal dilemmas, opposition and mistrust, logistical difficulties, and a lack of information and resources.<br>According to the study's findings, ChatGPT may provide important information to dispel COVID-19 vaccine conspiracies. It presents both arguments for and against mandatory vaccination in a straightforward, succinct, and impartial manner.<br>The study cautions that, despite ChatGPT's usefulness, content produced by it shouldn't take the place of trustworthy sources of vaccine information like the World Health Organisation (WHO) and the Centres for Disease Control and Prevention (CDC). |
| Salvagno et al 2023 | Italy | Review article | Peer reviewed | Can artificial intelligence help for scientific writing? | ChatGPT has demonstrated its usefulness in supporting researchers across various aspects of scientific writing, including drafting articles, abstracts, literature research, data summarization, structure suggestions, and language review.<br>However, there have been no published articles so far on the use of this technology in critical care medicine, leaving it uncertain whether its implementation would simplify or complicate the writing process within this discipline.<br>While ChatGPT cannot generate entirely new ideas, it can assist researchers by developing their existing ideas and creating initial drafts. It has the potential to aid in the review process by identifying and summarizing academic papers and highlighting areas of uncertainty.<br>During the writing process, ChatGPT can generate an initial draft of a scientific paper, suggest titles, justify sample sizes, and describe data analysis techniques.<br>Furthermore, ChatGPT can provide assistance in the editing process, including formatting, language editing, and summarizing the text to create a suitable abstract,<br>thereby saving researchers time.<br>However, it is important to carefully evaluate the quality of AI-generated content, as it cannot substitute human expertise, creativity, and critical thinking.<br>Future applications of AI may involve the automatic generation of figures, tables, and other visual elements for manuscripts, contributing to data summarization.<br>While AI can quickly comprehend information and establish connections between evidence, it lacks the nuance, style, and originality that human authors provide. It also carries the risk of perpetuating existing biases and inaccuracies present in the data.<br>Ethical concerns arise regarding AI-generated writing, including issues of plagiarism, bias, accuracy, and authorship.<br>In addition to scientific writing, ChatGPT has the potential to support physicians in their hospital work by providing information on recognized ICU protocols, generating clinical notes, and assisting in patient communication.<br>Despite its potential benefits, the technology's limitations must be recognized. The accuracy, current knowledge, and precision of the AI-generated content may be variable and should always be validated by human experts.<br>As AI and chatbot tools continue to develop, international academic regulations are needed to govern their use in scientific writing and establish mechanisms for identifying and penalizing unethical usage.<br>Ultimately, AI and chatbots like ChatGPT should not replace human researchers' expertise, judgment, and personality, but rather supplement them. |
| Sanmarchi et al 2023 | Italy | Original article | Preprint | A step-by-step Researcher's Guide to the use of an AI-based transformer in epidemiology: an exploratory analysis of ChatGPT using the STROBE checklist for observational studies | **Aim of the Study:** The purpose of this research was to evaluate how early-stage AI-based transformer models, specifically OpenAI's ChatGPT, can assist researchers in the multiple steps of an epidemiological study. The study was simulated using the STROBE framework and the responses of the transformer were assessed for coherence and relevance.<br>**Methods:** The study involved posing questions derived from the STROBE checklist to ChatGPT and assessing its answers in the context of conducting an epidemiological study. The answers were evaluated by three independent senior researchers. |
|  |  |  |  |  | STROBE Guidelines: The STROBE guidelines were used prospectively in the design and conduct of the study. These guidelines provide a framework for transparent reporting of observational studies, ensuring the quality and transparency of the research.<br>ChatGPT: ChatGPT is a third-generation, autoregressive language model by OpenAI that generates human-like text. It was used to provide support to the researchers conducting the epidemiological study.<br>Use of ChatGPT: ChatGPT was utilized by formulating questions from the STROBE recommendations and contextualizing them with a real epidemiological study. The STROBE recommendations were transformed into specific questions for ChatGPT, which then provided answers.<br>Expert Assessment: The outputs of the transformer were assessed through "human evaluation," where annotators were asked to rate the responses produced by the model on a 1 to 5 points Likert scale, taking into account the coherence and relevance of the responses.<br>Results: From the 35 STROBE recommendations, 35 specific questions were derived for ChatGPT. Three domain experts evaluated the answers. The overall mean coherence score was 3.6 out of 5.0, and for relevance, it was 3.3 out of 5.0. Some responses received high scores for coherence and relevance, while others scored poorly. |
| Sarink et al 2023 | The Netherlands | Letter to the editor | Peer reviewed | A Study on the Performance of ChatGPT in Infectious Diseases Clinical Consultation | An AI chatbot called ChatGPT employs deep learning to produce discourse that sounds human. By contrasting its performance with the recommendations of clinical microbiologists (CM) or ID specialists in 40 clinical consultations at a tertiary hospital in the Netherlands, its use in the field of infectious disease (ID) was investigated.<br>In the study, the output of ChatGPT was compared to the diagnosis or treatment recommendations made by the ID or CM specialists in the identical clinical settings. On a scale of 1 (bad, wrong advise) to 5 (outstanding, entirely correlating with the specialist's recommendations), the AI's output was evaluated.<br>The ChatGPT had a mean score of 2.8 and a standard deviation of 1.5. It performed best when interpreting positive blood culture results (mean 3.3, SD 1.3) and poorly when providing guidance on osteomyelitis or infections of prosthetic joints (mean 1.3, SD 0.5). These scores' differences were statistically significant.<br>ChatGPT occasionally supplied unclear treatment options and didn't always take into account given data. It occasionally made incorrect claims about the origin of its guidance, which raises questions about its veracity and accuracy. |
|  |  |  |  |  | ChatGPT's recommendations fully matched those of the experts in a few difficult situations, suggesting that it may be helpful in difficult circumstances.<br>The study's limitations include the exclusion of chronic cases and the possibility of differences between the data provided to ChatGPT and the reference standard clinical consultation.<br>The study comes to the conclusion that while ChatGPT can generate moderately accurate diagnostic or therapeutic suggestions, it cannot take the role of clinical expertise. Clinicians are required to direct and interpret ChatGPT's recommendations. |
| Schulte 2023 | USA | Original article | Peer reviewed | Capacity of ChatGPT to Identify Guideline-Based Treatments for Advanced Solid Tumors | This study aimed to evaluate the capability of ChatGPT in suggesting suitable systemic treatments for new cases of advanced solid malignancies, in comparison with the recommendations found in the National Comprehensive Cancer Network (NCCN) guidelines.<br>Researchers used standardized prompts to assess ChatGPT's ability and introduced a measurement called the valid therapy quotient (VTQ), which is a ratio of medications listed by ChatGPT to those recommended by the NCCN guidelines.<br>The study used 51 different diagnoses and ChatGPT was able to identify 91 different medications for the treatment of advanced solid tumors.<br>The overall VTQ was found to be 0.77, indicating a high level of concordance with the NCCN guidelines.<br>For each case, ChatGPT was able to provide at least one systemic therapy example that matched the NCCN's suggestions.<br>There was a weak correlation identified between the incidence of each malignancy and the VTQ.<br>The study concluded that ChatGPT showed significant ability in recognizing and suggesting medications used in the treatment of advanced solid tumors according to the NCCN guidelines.<br>However, the practical usefulness of ChatGPT in assisting oncologists and patients in deciding the treatment remains uncertain.<br>The study anticipates that future versions of ChatGPT will have improved accuracy and consistency in this area and emphasizes the need for further studies to quantify its capabilities more effectively. |
| Singh 2023 | India | Commentary | Peer reviewed | Artificial intelligence in the era of ChatGPT - Opportunities and challenges in mental health care | OpenAI launched ChatGPT, an AI-based chatbot, on November 30, 2022, and it has already amassed over 100 million users. Numerous other AI applications are on the horizon, including Google Bard, Microsoft Bing AI, Chinese Ernie bot, Korean SearchGPT, Russian YaLM 2.0, Chatsonic, Jasper Chat, Character AI, Perplexity AI, and YouChat.<br>AI platforms like ChatGPT are recognized for their vast potential across various domains, including mental health. They are being utilized for chatting, gaming, computer programming, and even generating scientific papers and abstracts.<br>There exists a substantial treatment gap in mental health care, particularly in developing and lower-income countries, where 76%-85% of mental disorders remain untreated. AI interfaces such as ChatGPT are viewed as potential tools to bridge this gap, providing companionship, support, and therapy for individuals with limited accessibility and affordability in mental health care.<br>However, while AI chatbots like ChatGPT have immense promise, there are concerns regarding their use in mental health. These concerns include the potential for providing incorrect information or advice, issues related to confidentiality, privacy, and data security, as well as challenges in accurately diagnosing mental health conditions.<br>Other concerns involve the lack of standardization, the possibility of misdiagnosis, the provision of inappropriate advice, and limitations in handling crises. These ethical concerns surrounding the use of AI-based applications in mental health are significant.<br>In response to these concerns, organizations like the American Psychiatric Association (APA) are establishing task forces to monitor and regulate AI-based mental health apps. The APA has developed an App Evaluation Model known as App Advisor, which is being adopted by other healthcare organizations for the assessment of health apps.<br>Given the significant variations in awareness, education, language, and understanding within the Indian population, the Indian Psychiatric Society and other stakeholders should begin evaluating and regulating AI-based global and local apps for their safety, efficacy, and tolerability. They should also provide guidance to the general public regarding proper and safe usage of these apps. |
| Singh et al 2023 | India | Original article | Peer reviewed | ChatGPT and Ophthalmology: Exploring Its Potential with Discharge Summaries and | he study examined the ability of the OpenAI model, ChatGPT, to generate operative notes and discharge summaries across various ophthalmic surgical specialties, maintaining the tenets of the Declaration of Helsinki. The surgical specialties included cataract surgery, penetrating keratoplasty, LASIK, trabeculectomy, pars plana vitrectomy, scleral buckle, pneumatic retinopexy, |
|  |  |  |  | Operative Notes | intraocular injections, open-globe injury repair, extraocular muscle surgery, Levator resection for ptosis, and dacryocystorhinostomy. The AI model's outputs were evaluated for their evidence-based content, specificity, factual accuracy, and ability to recognize mistakes.<br>Key findings from the study include:<br>The AI model could quickly generate detailed discharge summaries and operative notes in response to provided prompts, with responses usually returned in less than 20 seconds.<br>The generated summaries contained no disclaimers, indicating confidence in the provided information.<br>ChatGPT showed a capability to incorporate specific information such as medications, follow-up instructions, and consultation times into the discharge summaries when prompted.<br>While the AI model's summaries were generally accurate, they contained some generic text, particularly in the 'discharge instructions' section.<br>The operative notes were divided into several sections, including pre- and post-operative diagnosis, procedure, anesthesia, indication, procedure details, post-operative care, estimated blood loss, complications, and summary.<br>ChatGPT demonstrated a capacity to learn from its mistakes. When confronted with a potential error regarding the administration of sedation prior to intraocular injection of Ranibizumab, the AI model immediately apologized for the error and correctly generated the procedure without sedation when re-prompted.<br>Despite some shortcomings, the authors noted that the model's responses could be customized to the user's needs with further training. This study demonstrates the potential of AI in assisting with documentation tasks in the field of ophthalmology. |
| Tang et al 2023 | USA | Review | Preprint | Evaluating Large Language Models on Medical Evidence Summarization | This extensive study assesses the performance of large language models (LLMs) in medical evidence summarization using Cochrane Reviews across six clinical domains, including heart failure, kidney illness, esophageal cancer, Alzheimer's disease, and neurological problems.<br>The study compares the zero-shot performance of two models: GPT-3.5 and ChatGPT. These models were given different parts of the review abstracts to summarize. The models' output was assessed on several metrics including ROUGE-L, METEOR, BLEU, and manual human evaluation for factors like coherence, factual consistency, comprehensiveness, and harmfulness. |
|  |  |  |  |  | Findings indicate that all models perform similarly on automatic metrics with higher ROUGE scores showing that key information from the source document was effectively captured. However, the models were found to be more extractive compared to human-written summaries, indicating a lower level of abstraction.<br>Human tests revealed that ChatGPT's summaries were more cohesive than GPT3.5's. It is noteworthy that less than 10% of the summaries created by ChatGPT-MainResult included factual errors. In terms of thoroughness, ChatGPT-MainResult and ChatGPT-Abstract both delivered more than 75% of the time, with the former also producing the fewest summaries that were medically dangerous.<br>According to the survey, ChatGPT-MainResult performed much better than its competitors, delivering the most preferred summaries about half of the time among the three LLM setups. It was chosen because it produced the most thorough summary and contained more important information. |
| Temsah et al 2023 | Saudi Arabia | Original article | Peer reviewed | Overview of Early ChatGPT's Presence in Medical Literature: Insights From a Hybrid Literature Review by ChatGPT and Human Experts | Introduction: OpenAI's ChatGPT has emerged as a powerful AI model capable of generating human-like text in response to user queries, with applications spanning various domains, including medical education.<br>Methodology: This review utilizes a hybrid narrative methodology, combining traditional narrative review methods with the assistance of ChatGPT to analyze and synthesize abstracts.<br>Inclusion and Exclusion Criteria: Articles discussing ChatGPT within the context of medical education, literature, or practice were included. Non-medical or unrelated contexts, non-English sources, and articles published outside the specified date range were excluded.<br>Results: A total of 65 papers were identified in PubMed and 110 papers in Europe PMC. These papers highlighted the utilization of ChatGPT in diverse medical fields such as medical education, scientific writing, research, and diagnostic decision-making.<br>Key Findings: The review identified eight main themes: (1) medical writing, (2) medical education, (3) diagnostic decision-making, (4) public health, (5) scientific research, (6) ethical considerations of ChatGPT use, (7) ChatGPT's potential to automate medical tasks, and (8) criticism of ChatGPT's usage.<br>ChatGPT in Medical Writing: ChatGPT demonstrates the ability to generate coherent and readable content, but concerns arise regarding its accuracy and the ethical implications it introduces. |
|  |  |  |  |  | ChatGPT in Medical Education: ChatGPT has the potential to enhance learning, interpretation, and recall of medical information, but concerns persist regarding the undermining of clinical reasoning and the lack of context or tailoring to individual learning needs.<br>ChatGPT in Diagnostic Decision-Making: ChatGPT can improve efficiency and reduce errors in diagnostic decision-making, but concerns exist regarding accuracy, potential bias, and the need for human oversight.<br>ChatGPT in Public Health: ChatGPT can provide reliable estimates in public health research, provided the input data is valid and accurate.<br>ChatGPT in Science and Research: It can streamline scientific writing processes and assist with literature reviews, data interpretation, and hypothesis generation. However, concerns are raised regarding its accuracy and potential to undermine human expertise.<br>Ethical Considerations of ChatGPT: It raises concerns about authorship, accountability, and transparency, along with the potential for generating misleading or inaccurate information.<br>ChatGPT's Potential to Automate Medical Tasks: It can potentially improve efficiency and speed of programming but undermines the value of human expertise.<br>Criticism of ChatGPT Usage: The model's limitations, such as the production of erroneous content, susceptibility to bias, and need for stringent regulatory safeguards, are discussed.<br>Overall, this review highlights the transformative potential of ChatGPT. |
| Thorp 2023 | USA | Commentary | Peer reviewed | ChatGPT is fun, but not an author | ChatGPT, developed by OpenAI, has become a cultural sensation within two months of its release, providing endless entertainment by creating text based on written prompts.<br>Despite its popularity, the AI has been noted to sometimes write plausible but incorrect or nonsensical answers. This includes referencing non-existent scientific studies.<br>Concerns have been raised about the impact of ChatGPT on education, as it can write essays on a range of topics. While the program can generate factual answers, the quality of scholarly writing is currently lacking.<br>The emergence of AI tools like ChatGPT is prompting academics to rethink their course designs and assignments to ensure they aren't easily solvable by AI. |
|  |  |  |  |  | Concerns also exist regarding the influence of AI on scientific paper writing. In a study, only 63% of abstracts created by ChatGPT were identified as AI-generated by academic reviewers.<br>Science journals are now updating their licenses and editorial policies to explicitly state that text generated by AI tools, such as ChatGPT, cannot be used in their work. This includes figures, images, or graphics produced by AI tools.<br>Violating these policies will be considered scientific misconduct, akin to plagiarism or altering images.<br>The journals recognize that most instances of scientific misconduct occur due to lack of adequate human attention, hence the need for scientists to be vigilant in their work.<br>The role of AI should ultimately be as a tool to aid people in posing hypotheses, designing experiments, and making sense of results in the scientific process. |
| Haq et al 2023 | Pakistan | Original article | Preprint | Comparing human and artificial intelligence in writing for health journals: an exploratory study | The study aimed to investigate the suitability of ChatGPT for scientific writing by comparing short articles written by human authors and ChatGPT on three different topics.<br>The research included criteria for testing ChatGPT's use in scientific writing and developed evaluation criteria to assess the quality of articles written by human authors and ChatGPT.<br>The evaluation criteria were adapted from the EASE guidelines, focusing on three major areas: structure, scientific content, and credibility.<br>Grammarly, another AI-based tool, was used to score two items: originality and readability.<br>The team selected three diverse topics for human authors and ChatGPT to write about, shared the output as blinded versions, and evaluators scored the articles independently.<br>Results showed that while human-authored articles scored perfectly, ChatGPT articles lacked in areas like structure and organization, did not follow the IMRaD (Introduction, Methods, Results, and Discussion) structure and lacked a logical flow.<br>In terms of credibility, human-authored articles again scored a perfect five, but ChatGPT articles had modest scores. Citations in ChatGPT articles were minimal<br>and often pointed to nonexistent resources.<br>On scientific content, both types of articles scored well on originality, but ChatGPT articles were weaker in specificity of response, lack of numerical data, cohesiveness, and inclusion of study limitations.<br>Efficiency was the only area where ChatGPT scored perfectly, taking significantly less time to produce the articles compared to human authors.<br>The study concluded that while ChatGPT can efficiently produce original and seemingly coherent articles, they lack in-depth scientific basis, lack reproducibility, and might quote non-existent scholarly work. This raises questions about the accuracy and reliability of ChatGPT in the context of scientific writing. |
| Uprety et al 2023 | USA | Review article | Peer reviewed | ChatGPT-A promising generative AI tool and its implications for cancer care | ChatGPT is a transformative AI model developed by OpenAI, offering sophisticated, human-like text-based conversations. With the release of GPT-4, ChatGPT Plus has seen widespread interest and potential applications, particularly within the healthcare sector.<br>ChatGPT can be utilized in the medical field for extracting crucial information from patient records, saving physicians time and enhancing efficiency. This is particularly beneficial in areas such as oncology, where patients often have extensive and complicated histories.<br>The AI can be used for administrative tasks, such as creating insurance letters for evidence-based authorization of therapies.<br>ChatGPT could aid in the rapidly evolving field of oncology by staying updated on recent advances and approvals, aiding in the interpretation of complex Next-Generation Sequencing (NGS) reports, and making appropriate treatment recommendations based on the correlation between mutations (biomarkers) and treatment drugs.<br>The AI can support oncologists in making suitable clinical trial recommendations for patients by interpreting their detailed medical history and the range of content available on ClinicalTrials.gov.<br>Despite its vast potential, ChatGPT currently has some limitations. It's not compliant with the Health Insurance Portability and Accountability Act (HIPAA), posing a risk of violating patient privacy. Additionally, the AI could unintentionally propagate biases and misinformation inherent in the internet text it was trained on, and it is vulnerable to adversaries and manipulation.<br>ChatGPT plugins could enhance cancer care by efficiently summarizing medical<br>documentation, interpreting NGS testing reports, and suggesting relevant clinical trials for patients. However, it needs to overcome its current limitations related to HIPAA compliance, internet bias, and vulnerability to adversarial prompting.<br>The authors anticipate that future iterations of ChatGPT may overcome these limitations and become a highly valuable tool for healthcare, particularly in the field of oncology. |
| Uz and Umay<br>2023 | Turkey. | Original<br>article | Peer<br>reviewed | "Dr ChatGPT": Is it a<br>reliable and useful<br>source for common<br>rheumatic diseases? | The objective of this study was to utilize Google Trends to identify the most frequently searched terms related to common rheumatic disorders (OA, RA, FMS, AS, SLE, gout, and PSA). Based on the search engine results, keywords were determined and categorized for each condition. Likert-type ratings were employed to evaluate the accuracy and value of ChatGPT's responses to these keywords.<br>The most commonly searched keywords related to OA included "knee osteoarthritis," "osteoarthritis pain," "osteoarthritis hip," and "osteoarthritis treatment." For RA, popular terms were "rheumatoid arthritis symptoms," "rheumatoid arthritis pain," "rheumatoid arthritis causes," and "rheumatoid arthritis treatment."<br>AS-related keywords encompassed "ankylosing spondylitis pain," "ankylosing spondylitis symptoms," "ankylosing spondylitis test," and "ankylosing spondylitis treatment." SLE was associated with keywords such as "lupus disease," "lupus symptoms," "lupus causes," and "lupus treatment." PSA keywords included "psoriatic arthritis symptoms," "psoriatic arthritis pain," "psoriatic arthritis causes," and "psoriatic arthritis treatment." FMS was linked to terms like "what is fibromyalgia," "fibromyalgia pain," "fibromyalgia symptoms," and "fibromyalgia treatment."<br>Using Likert-type scales, the study evaluated the accuracy and value of ChatGPT's responses for each keyword. The reliability and usefulness scores demonstrated good to very good agreement, as indicated by Cronbach values and inter-rater reliability scores.<br>Overall, the dependability and usefulness scores ranged from 4 to 7. AS received the highest usefulness score, while OA received the highest reliability score. Some topics received lower marks in terms of usefulness and reliability. However, there were no significant discrepancies in the ratings for overall dependability and usefulness among the raters.<br>This study comprehensively examined the accuracy and value of ChatGPT's responses to the most popular search terms related to common rheumatic conditions. The findings provide insights into the precision and applicability of AI-generated knowledge for individuals seeking information about these disorders. |
| Van Dis et al 2023 | the Netherlands | Commentary | Peer-reviewed | ChatGPT: five priorities for research | Large Language Models (LLMs) like ChatGPT have the potential to revolutionize the scientific research field, offering assistance in various tasks including writing essays, summarizing literature, and identifying research gaps.<br>However, there is controversy around this technology due to its capacity to produce convincingly wrong information, potentially distorting scientific facts and spreading misinformation.<br>LLMs can introduce inaccuracies, bias, and plagiarism. ChatGPT, for instance, has been found to generate false and misleading responses when asked complex, nuanced scientific questions.<br>Like humans, current AI models are susceptible to biases, such as availability, selection, and confirmation biases, which they can inadvertently propagate.<br>Researchers using AI like ChatGPT risk being misled by false or biased information and incorporating it into their work. Unintentional plagiarism is also a concern as AI reproduces text without reliable citations.<br>The researchers suggest the necessity of fact-checking and verification processes, even with AI assistance, to maintain the integrity of scientific practice.<br>Accountability rules for AI usage in research are proposed, including transparency in acknowledging AI assistance in papers and the development of policies to handle the use of AI in manuscript preparation.<br>There are emerging questions about authorship and rights to texts created with AI assistance, indicating the need for updated definitions and legal considerations.<br>The proprietary nature of AI technologies, mostly controlled by a few large tech companies, raises ethical concerns and hampers transparency. Open-source, independent non-profit projects for AI development are recommended.<br>Despite the challenges, the potential benefits of AI in reducing workload, speeding up publication, and potentially accelerating innovation are recognized. Investment in the validity and reliability of LLMs for effective use in scientific research is suggested.<br>AI could change the academic skill set, optimizing training, reducing the need for certain skills, and introducing new ones. As AI advances, it might handle more complex tasks, prompting the need for careful evaluation of AI acceleration vs loss of human potential. |
|  |  |  |  |  | The research community is urged to engage in a wide-ranging debate on the use and implications of LLMs in research, with focus on topics like essential academic skills, steps requiring human verification, and ensuring LLMs promote equity in research.<br>Every research group should discuss the use and implications of LLMs, and educators should introduce discussions about LLM usage and ethics to students. Accountability for research work will apply regardless of whether it's generated with ChatGPT or not. |
| Waisberg et al 2023 | Ireland | Letter to the editor | Peer reviewed | GPT-4: a new era of artificial intelligence in medicine | A sizable language model called ChatGPT was created by OpenAI. To anticipate words or phrases in context, it employs a deep neural network based on the Transformer architecture.<br>The model is trained to anticipate words in a sentence or phrase depending on words that came before them. Recursively repeating the technique until a whole sentence or paragraph is generated will accomplish this.<br>ChatGPT recognises links between words and sentences in natural language to deliver logical and pertinent responses to user inputs.<br>The AI processes user input, analyses it, and then responds in accordance with the patterns it identified during training. This response is intended to be situationally suitable and can cover a wide range of issues.<br>Banking, gaming, and healthcare are just a few of the industries where artificial intelligence is being used. It might help with patient diagnosis, treatment alternatives, and individualised care plans, for instance, in the healthcare industry.<br>The AI was instructed to write an easy-to-read discharge narrative for a patient who had undergone a simple cholecystectomy. Important recommendations on exercise limitations, dietary modifications, and when to seek medical assistance were all included in the summary.<br>The AI was also asked to provide information about the latest clinical trials to treat interstitial lung disease (ILD). It listed four ongoing phase 2 clinical trials and provided brief details about each one.<br>Furthermore, the AI was asked to provide the latest guidelines for AI in medicine. It listed guidelines from the European Commission, the American Medical Association, the International Medical Informatics Association, and the National Institute for Health and Care Excellence, all of which were relevant. |
|  |  |  |  |  | In contrast, the AI's newly implemented image analysis feature demonstrated a lack of accuracy in identifying fundus photographs of anterior ischemic optic neuropathy and non-arteritic anterior ischemic optic neuropathy. Instead, it erroneously classified these images as a schematic representation of the CRISPR-Cas system.<br>Despite the inaccurate image analysis, GPT-4 is considered an improvement over its predecessor, with enhanced problem-solving abilities, an expanded knowledge base, and the ability to process eight times more words. It's also more difficult to deceive and is less likely to respond to inappropriate requests. |
| Wen and Wang 2023 | Australia | Commentary | Peer-reviewed | The future of ChatGPT in academic research and publishing: A commentary for clinical and translational medicine | ChatGPT in Medical Research: ChatGPT, a large AI model developed by OpenAI, is being increasingly used in various occupations including clinical and translational medicine. It has entered the scientific literature with published papers and preprints, however, its limitations in medical research, particularly in providing accurate and up-to-date information, are causing concern.<br>Factual Inaccuracies of ChatGPT: The biggest disadvantage of ChatGPT is its potential to compile inaccurate information. It uses data up until 2021, and does not consider information after that, which can be a hindrance in fields like medicine where knowledge and advances are constantly evolving.<br>ChatGPT: Difficult to Detect: Researchers at Northwestern University conducted a study where ChatGPT was asked to write 50 medical research abstracts. Medical researchers had difficulty distinguishing between AI-generated and human-written abstracts, raising concerns about the potential consequences of relying on potentially inaccurate research.<br>ChatGPT as an Aide for Scientific Innovation: Despite its limitations, ChatGPT can be used beneficially in scientific research to improve completed research papers. It can increase researchers' productivity, save time, and improve the quality of their content.<br>Pros and Cons of ChatGPT in Scientific Research and Publishing: While ChatGPT can be useful for certain tasks like editing, it is not reliable for providing accurate facts or references. Using it judiciously while being aware of its limitations can help researchers streamline their work without risking the publication of false information.<br>ChatGPT and Clinical and Translational Medicine: Implementing ChatGPT must be pursued cautiously due to its limitations in providing reliable information. It can be useful for tasks like proofreading and manuscript checks, but cannot replace the specialized knowledge, innovative ideas, and opinions that human input provides. Further exploration of research ethics and the moral use of AI in health research is required to establish guidelines. |
| Xue et al 2023 | China | Commentary | Peer-reviewed | The potential impact of ChatGPT in clinical and translational medicine | ChatGPT is an AI model developed by OpenAI that simulates human interaction and uses a deep learning technique called 'transformer architecture'. It's trained on large datasets of text from the internet.<br>It is still unclear how ChatGPT will affect clinical medicine. However, similar AI models could help with patient education, clinical trial recruiting, clinical data management, research support, clinical decision support, and the automation of activities like data processing and picture interpretation.<br>ChatGPT could also assist in drug discovery by recognizing, classifying, and describing chemical formulas or molecular structures. It can also help in disease prediction, diagnosis, and assessment of therapeutic targets.<br>ChatGPT has restrictions despite its advantages. It occasionally gives general or ambiguous responses in conversations about medicine since it is unable to update its training data in real-time. ChatGPT's diagnoses could not be thorough or sufficient, showing that it is still unable to manage the challenging tasks of clinical practise on its own.<br>It has been claimed that emphasising ChatGPT's use in human-computer interaction could increase its usefulness in clinical settings. By speeding data gathering, recording, and analysis, it could assist with patient questionnaires and interviews in epidemiological research as well as mental health care.<br>ChatGPT has already been used in medical education, training, and writing, though ethical issues around its use have arisen, particularly regarding its authorship of academic articles.<br>AI models, including ChatGPT, can aid healthcare by providing a more objective, evidence-based approach to decision-making and by helping to identify patterns and correlations in large datasets. They can also assist in disease detection and prognosis prediction.<br>AI may have unfavourable effects on society, such as privacy problems and bias or discrimination. To ensure the responsible and effective use of AI in healthcare, more research and development is required.<br>Even so, ChatGPT and other forms of AI will continue to advance. The best course of action is to embrace it, making use of its potential to advance clinical practise while simultaneously addressing any potential drawbacks. |
| Yadava 2023 | India | Editorial | Peer-reviewed | ChatGPT—a foe or an ally? | ChatGPT, an AI platform developed by OpenAI, employs natural language processing technology, using advanced machine learning paradigms such as transfer learning, supervised learning, and reinforced learning. |
|  |  |  |  |  | The model has the capability to generate articulate and meaningful content across various fields of knowledge, with outputs including music, plays, poetry, and song lyrics.<br>Despite its advanced abilities, there are concerns within academia about the impact of non-human authors on the integrity and validity of scientific publications. This could potentially lead to an increase in academic plagiarism.<br>Some renowned scientific journals such as 'Nature' and 'JAMA Network Science' have decided not to accept articles generated by ChatGPT, demanding full disclosure of its use.<br>The technology has also sparked worries among non-medical intelligentsia and politicians, with fears about potential job losses, discrimination, and uncontrollable military applications.<br>Historical examples show that disruptive technologies often face resistance upon introduction. However, despite initial skepticism, many have gone on to have profound impacts on society and industry.<br>Some academic journals and researchers have already adopted the technology, recognizing its potential benefits for medical writing and document creation.<br>The technology is expected to streamline scientific writing and administrative tasks, improving efficiency in clinical roles.<br>The debate on the implications of AI like ChatGPT is ongoing. It could turn out to be a dual-use technology, where the ethics and intent behind its use determine its value.<br>Regardless of the pros and cons, AI-powered language models are becoming a reality, with human oversight remaining crucial.<br>The adoption of AI technology is seen as inevitable, and it is essential to accept, adapt, and leverage it for the future. |
| Yeung et al 2023 | UK | Commentary | Peer-reviewed | AI chatbots not yet ready for clinical use | A Cambrian explosion of natural language processing (NLP) models, predominantly based on the transformer model, occurred in 2022, offering unprecedented advancements in natural language generation capabilities and opening up the potential for their application in healthcare as AI chatbots.<br>Large Language Models (LLMs), like OpenAI's GPT3 and Google's PALM, Gopher, and Chinchilla, have evolved to exhibit emergent properties: performing tasks they<br>were not explicitly trained on. This could be due to the models' ability to extract more knowledge from vast amounts of text data used in their training.<br>LLMs have shown promise in the healthcare field, exhibiting human-level performances in medical question answering and summarisation tasks. However, safety and accuracy should be prioritised over human-like interactivity.<br>Current LLMs face challenges such as mirroring biases and inaccuracies from their training data, raising concerns about their suitability for critical fields like healthcare. Moreover, LLMs trained on biomedical corpora, while being domain-specific, don't necessarily reflect the realities of actual patients and diseases in healthcare.<br>Some LLMs, such as GatorTron and BEHRT, are trained on de-identified clinical notes or disease classification codes to circumvent the problem of sensitive patient data.<br>Current medical AI benchmarks do not adequately capture the complexity of real-world clinical practice. In actual practice, healthcare professionals generate a list of potential differential diagnoses and then proceed with investigations to gradually narrow down the possibilities, rather than simply identifying a single correct diagnosis.<br>In a comparative study between ChatGPT and Foresight GPT (trained on real-world hospital data), both models performed well in predicting the 5 most likely diagnoses from synthetic clinical histories. However, ChatGPT, which is not domain-specific, often omitted crucial diagnoses.<br>LLMs can exhibit biases and associations based on their training text, leading to potential issues of racial-ethnic disparities in treatment recommendations.<br>LLMs can also generate "hallucinations" or false information, especially when faced with insufficient or misleading information in the prompt.<br>Although LLMs have made encouraging strides in natural language processing and creation, it is still unclear whether they are ready to be used as clinical tools that patients will see. Focus should be placed on domain-specific training data, expert clinician fine-tuning, and transparent depiction of output relevancy versus safety implications in order to use AI chatbots in healthcare in a safe manner. |
| Young et al 2023 | USA | Research Letter | Peer reviewed | The utility of ChatGPT in generating patient-facing and clinical responses for melanoma | The study aimed to assess the effectiveness of OpenAI's ChatGPT, a large language model (LLM), as an informational platform for patients seeking information about melanoma care. To evaluate its suitability, twenty-five hypothetical patient questions related to melanoma were presented to three board-certified dermatologists. The dermatologists assessed the appropriateness of the<br>responses for a patient-facing informational platform, their sufficiency for clinical practice, accuracy, and comprehensibility.<br>The accuracy of ChatGPT's responses consistently received high ratings, with an average score of 4.88 out of 5. Furthermore, 92% of the responses were considered appropriate for a patient-facing informational platform.<br>However, the study found that only 64% of the responses were deemed sufficient for clinical practice. This was mainly due to important details being omitted in ChatGPT's advice. For instance, while recommending "regular skin exams" is accurate, it lacks the specificity of a physician's recommendation for exams every few months.<br>Another limitation highlighted by the study was the average readability of ChatGPT's responses, as measured by the Flesch Reading Ease Score (FRES). The FRES indicated a college-level comprehension requirement, which is too advanced for the general public. Health-related materials are typically recommended to be at a 5th-6th grade reading level.<br>In conclusion, the study acknowledges that ChatGPT can generate accurate responses suitable for patient-facing tools. However, it emphasizes significant limitations, particularly in terms of comprehensibility and sufficiency for clinical practice. The authors stress the importance of dermatology providers understanding the potential uses and limitations of AI tools like ChatGPT to effectively counsel patients as these technologies become more prevalent in clinical practice. |
| Zheng and Zhan 2023 | USA | Commentary | Peer reviewed | ChatGPT in Scientific Writing: A Cautionary Tale | ChatGPT is an AI chatbot that has garnered worldwide excitement and concerns, particularly in the scientific community due to its use in scientific writing and publishing. The concerns are mainly related to copyright, attribution, plagiarism, authorship, and the accuracy of the content generated by ChatGPT.<br>The authors conducted an evaluation of ChatGPT's accuracy in generating content by using an article on Body Surface Area (BSA) formulas that was not included in its training data. They summarized key facts from the article and used them as prompts to ask ChatGPT questions five times, assessing the responses for accuracy.<br>While ChatGPT generates responses that are well-written and sound plausible, it has been observed that these responses often contain significant errors and fabricated information. Consequently, ChatGPT cannot be considered a reliable and trustworthy source for scientific writing.<br>Given the potential for falsifications and fabrications in ChatGPT's output, the study<br>concludes that there is no clear advantage to using ChatGPT for scientific writing. Authors utilizing ChatGPT must take the responsibility to manually verify all facts, statements, and references generated by the AI.<br>Upholding the highest ethical standards in scientific research is paramount, and proper data management is essential in maintaining these standards. Poor data management practices, including the presence of fabricated or falsified findings, can have serious consequences.<br>It is important to note that the authors listed on an article are ultimately responsible for ensuring accuracy and integrity, not ChatGPT itself. Detecting fabrication or falsification during the peer-review process of manuscripts containing text generated by ChatGPT is challenging. Studies have shown that human reviewers can overlook up to 32% of fully fabricated abstracts produced by ChatGPT.<br>Directly adopting full text written by ChatGPT may constitute plagiarism and violate the code of conduct in scientific publishing.<br>Scientific journals that accept articles involving ChatGPT in the writing process may face a significant increase in retractions and loss of credibility.<br>While ChatGPT offers great potential for the future, its current state is not mature enough for scientific writing. The role of a more advanced ChatGPT in scientific writing necessitates comprehensive discussions and debates.<br>Considering the potential for fabricated and inaccurate information, the Science family of journals has implemented a ban on all ChatGPT-generated content. It is recommended that this policy becomes standard practice for all scientific publishing.<br>Despite its limitations, ChatGPT can still be a useful tool for checking grammar and syntax errors, as well as refining language, particularly for non-native speakers. |
| Zhong et al 2023 | China | Commentary | Peer reviewed | The Artificial intelligence large language models and neuropsychiatry practice and research ethic | Artificial intelligence (AI) can significantly enhance psychiatric research and practice by improving diagnostic accuracy, optimizing treatment outcomes, and offering personalized care. AI can analyze extensive patient data, recognize complex patterns, and suggest individualized treatment strategies.<br>AI large language models (LLMs) like ChatGPT, despite their potential benefits, raise concerns such as reliability and accuracy, transparency, accountability, and ethical implications. These models can sometimes generate plausible yet incorrect or nonsensical information, leading to doubts about their scientific reliability. |
|  |  |  |  |  | The transparency of AI LLMs is a challenge due to the complex algorithms and large datasets they employ. The opaque nature of these models could potentially undermine trust and credibility in scientific research where transparency is vital.<br>Accountability is another issue with AI LLMs. They could perpetuate biases present in the training datasets, and this could lead to biased outputs. For instance, if the AI model is trained on racially or ethnically biased data, its outputs may reflect these biases.<br>Ethical concerns with AI in research include the potential for these models to generate misleading content. Some cases have reported AI-generated abstracts that fooled academic reviewers. AI LLMs' increasing usage for tasks like data analysis, literature reviews, grant proposals, etc., is stirring debate about whether they should be acknowledged as authors in research papers.<br>There are concerns over intellectual property rights violations as AI models are trained on vast datasets that may include copyrighted or proprietary content. There's a risk that these models could generate content that infringes on these rights.<br>The article suggests measures to address these concerns and regulate AI LLMs use in science. These include insisting on human review of AI-generated content, establishing accountability mechanisms, investing in open-source LLMs, and promoting a broad discussion among stakeholders on the implications and challenges of AI.<br>Despite the potential issues, AI can be transformative in psychiatric research and practice. However, it's essential to address these concerns and ensure that AI's use is ethical, protects patients, and maximizes its benefits. |
| Zhou et al 2023 | China | Letter to the editor | Peer reviewed | The Potential of Applying ChatGPT to Extract Keywords of Medical Literature in 5 Plastic Surgery | Summary could not be generated. |
| Zhou 2023 | China | Original article | Peer reviewed | Evaluation of ChatGPT's Capabilities in Medical Report Generation | The study explores the potential of ChatGPT, an OpenAI language model, in supporting healthcare professionals by generating medical reports based on real patient laboratory results.<br>The language model was employed in various medical domains, such as interpreting lab results and analyzing medical literature, to improve and expedite the process of writing medical reports.<br>In this particular case, ChatGPT produced a detailed medical report for a 31-year-<br>old male patient who presented with abdominal pain and sought medical attention.<br>Based on routine laboratory tests, the model generated personalized recommendations for the patient, including suggested lifestyle modifications and potential medical treatment options.<br>Additionally, the model advised the patient to seek a gastroenterologist's consultation for further assessment and consideration of advanced treatment plans.<br>The structure and organization of the case report were entirely generated by ChatGPT, utilizing the patient's physical information and lab results as input.<br>To validate the accuracy and reliability of ChatGPT's recommendations, the generated report will be compared with an online doctor consultation system.<br>The study aims to demonstrate that ChatGPT can consistently and accurately produce coherent, comprehensive, and clinically relevant medical reports, showcasing its potential in aiding healthcare professionals. |
| Zhu et al 2023 | China | Commentary | Peer reviewed | Can the ChatGPT and other Large Language Models with internet-connected database solve the questions and concerns of patient with prostate cancer? | The researchers conducted an evaluation of multiple large language models (LLMs), including ChatGPT, YouChat, NeevaAI, Perplexity, and Chatsonic, to assess their usefulness in providing accurate and comprehensive information about prostate cancer (PCa).<br>To evaluate the LLMs' performance, the researchers designed 22 questions based on patient education guidelines and their own clinical experience. They assessed the LLMs in terms of accuracy, comprehensiveness, patient readability, humanistic care, and stability.<br>Most of the LLMs exhibited accuracy rates above 90%, with ChatGPT demonstrating the highest accuracy. Interestingly, the free version of ChatGPT performed slightly better than the paid version.<br>The LLMs generally provided comprehensive responses, addressing different aspects such as the significance of PSA levels, detailed treatment comparisons, and advising patients to consult their doctors.<br>In terms of readability, most LLM responses were satisfactory and displayed humanistic care, particularly when discussing PCa's relatively long survival time. However, not all inquiries received the same level of humanistic care.<br>One issue identified was the inclusion of outdated or incorrect information in some LLM responses. Examples include inaccurate comparisons between apalutamide and enzalutamide, and claims that open surgery was more common than robot-<br>assisted surgery for radical prostatectomy.<br>NeevaAI, in particular, tended to simply relay literature content without summarizing or explaining, resulting in poor readability. The study also found that real-time internet-connected LLMs did not outperform ChatGPT, which had limited access to data. This suggests that model training may be more crucial than real-time internet connection.<br>Despite their imperfections, LLMs show promise in providing accurate basic information about PCa. They could potentially be applied in patient education and consultation, facilitating shared decision-making and democratizing medical knowledge.<br>However, the researchers caution against replacing doctors with LLMs at this stage. LLMs may contain errors, omit important points, and struggle to analyze specific contexts. They also lack the ability to ask follow-up questions for further information or provide the same level of comfort as a human healthcare provider. |
| Zielinski et al 2023 | UK | Commentary | Peer reviewed | Chatbots, ChatGPT, and Scholarly Manuscripts: WAME Recommendations on ChatGPT and Chatbots in Relation to Scholarly Publications | Chatbots, including ChatGPT, are AI tools used in diverse fields, including healthcare, customer service, and education. They can create new content by processing and reorganizing existing information.<br>Despite its potential, ChatGPT has several limitations. It can generate incorrect or nonsensical answers and it may fail to ask clarifying questions. Its knowledge is restricted to what it learned before 2021.<br>OpenAI is working on improving ChatGPT. Other companies are also developing similar generative AI tools.<br>Chatbots can only generate output based on their training data and they cannot produce truly original thoughts. They may unintentionally plagiarize from their training materials.<br>ChatGPT can produce false statements, though it does not possess the intentionality to lie in the human sense.<br>As software tools, chatbots cannot be held legally responsible for their output. Liability falls on the users.<br>Including ChatGPT as an author is controversial and potentially legally indefensible.<br>ChatGPT represents a threat to scholarly journals due to potential introduction of false or plagiarized content. Peer review may fail to identify AI-generated content. |
|  |  |  |  |  | AI tools like DALL-E 2 and Imagen, which generate images, have similar concerns to ChatGPT. The provenance of images should be clearly indicated.<br>Editors need to establish policies on the use of AI tools like ChatGPT and require tools for detecting AI-generated content.<br>Recommendations include disqualifying chatbots from authorship, requiring transparency when chatbots are used, ensuring authors take responsibility for AI-generated content, and providing editors with AI-detection tools. |
| Zimmerman 2023 | USA | Commentary | Peer reviewed | A Ghostwriter for the Masses: ChatGPT and the Future of Writing | Natural Language Processing (NLP) is a subfield of computer science that enables computers to understand, respond to, and mimic human language in text form. This technology has been instrumental in developing models such as ChatGPT.<br>ChatGPT is a product of two-phase training: unsupervised pre-training on a large dataset and supervised fine-tuning on smaller datasets. This results in an effective model able to provide high-quality responses to diverse tasks and prompts.<br>NLP is already used in various applications like Siri, Alexa, Grammarly, and even in automated journalism. In the medical field, NLP is used for tasks such as information extraction, productivity improvement, and documentation.<br>ChatGPT, a product of OpenAI, has gained significant attention due to its ability to generate human-like content, ranging from passing scores on the USMLE exam to generating convincing medical abstracts.<br>While tools like Wordtune and Paperpal assist with sentence restructuring, ChatGPT can help restructure entire manuscripts and provide feedback on limitations.<br>ChatGPT is leading in its space, but it is not without competitors. Companies like Google, Facebook, and Anthropic are working to create models with similar capabilities.<br>Despite its impressive capabilities, ChatGPT has been met with criticism, including the potential for generating confabulatory outputs (hallucinations) when it does not have an appropriate response and potential bias.<br>Ethical considerations are also paramount when it comes to the originality of the content produced by these tools, making transparency of use crucial.<br>Corporate investment in AI systems has been growing over the years, indicating a growing interest and trust in the technology. However, the responsibility of using<br>these tools ethically and responsibly lies with the user. |

**Table-2:** Summaries of full text of 118 selected article were uploaded in pdf format on ChatPDF, subsequently 12 questions were asked. Questions and respective answers given by ChatPDF are listed in this table.

| Number | Question | Answer by ChatPDF | Summary |
| --- | --- | --- | --- |
| 1 | **What are potential role of ChatGPT in medical writing and research?** | ChatGPT can be used in medical writing to produce coherent and readable content. It can also be used in scientific research to generate creative ideas, write essays and homework assignments, and even contribute as an author on research papers. Additionally, ChatGPT has the potential to automate medical tasks and enhance learning, interpretation, and recall of medical information. However, concerns persist about its accuracy, potential biases in the training data, privacy concerns, issues related to authorship in academic papers, and ethical considerations of its use. Despite these challenges, AI tools like ChatGPT are expected to be widely adopted in the medical field in the future. | **ChatGPT has applications in medical writing, scientific research, and automation of medical tasks, but concerns remain regarding accuracy, biases, privacy, authorship, and ethics. Adoption in the medical field is anticipated.** |
| 2 | **What could be the role of ChatGPT in clinical practice?** | ChatGPT can assist with patient inquiries, help with writing medical notes and discharge summaries, aid in decision-making and treatment planning, and support medical educators in various tasks. In clinical practice, ChatGPT could potentially be used as a valuable tool for personalized learning and encouraging a shift towards critical thinking | **ChatGPT offers assistance in patient inquiries, medical note writing, decision-** |
|  |  | and problem-based learning. However, it is important to note that there are challenges associated with the use of ChatGPT in clinical practice, such as the need for access to current information, potential biases in the training data, privacy concerns, issues related to authorship in academic papers, and ethical considerations of its use. Therefore, regulations and control mechanisms should be established to address ethical utilization of ChatGPT and similar tools in medicine. | **making, and medical education, but challenges like biases, privacy, authorship, and ethics need to be addressed for ethical utilization in medicine.** |
| 3 | **What are ethical issues associated with paper writing?** | Paper writing entails several ethical concerns, including the risks of bias, plagiarism, copyright infringement, transparency issues, legal implications, lack of originality, incorrect responses, limited knowledge, and inaccurate citations. These concerns arise from the potential for ChatGPT to generate fabricated and unreliable research outputs, as well as the lack of transparency and accountability in its responses. Addressing these ethical issues is crucial to uphold research integrity and ensure the reliability and trustworthiness of the generated results. | **Ethical concerns in paper writing with ChatGPT involve bias, plagiarism, transparency, legal issues, and lack of accountability, which need addressing for reliable and trustworthy research.** |
| 4 | **Can CHATGPT be an author?** | The question of whether ChatGPT can be considered an author is a subject of controversy and may pose challenges in a legal context. The traditional authorship criteria, such as those outlined by the International Committee of Medical Journal Editors (ICMJE), require individuals to contribute substantially to various aspects of the research process, including conception, design, analysis, interpretation, and manuscript preparation. As an AI language model, ChatGPT cannot independently fulfill these criteria. | **Including ChatGPT as an author is controversial due to its inability to meet authorship criteria, but it can be used as a tool** |
|  |  | However, ChatGPT can serve as a valuable tool to support human authors in writing papers and generating ideas. Its contributions can be acknowledged in the paper, but it may not meet the requirements for authorship in the traditional sense.<br>Determining whether or not to include ChatGPT as an author should be approached on a case-by-case basis and aligned with established ethical guidelines for scientific research. It is essential to consider the specific contributions made by ChatGPT and ensure that human authors take responsibility for the substantial aspects of the research process as outlined by the ICMJE guidelines. | **to assist human authors. Decisions should be made on a case-by-case basis, adhering to ethical guidelines.** |
| 5 | **Can CHATGPT write text in good English and free of plagiarism?** | ChatGPT can assist in the editing process, including language editing, and summarizing the text to compose a suitable abstract. However, the quality of the AI-generated content needs careful evaluation as it is not a substitute for human expertise, creativity, and critical thinking. While ChatGPT can rapidly comprehend information and connect evidence, it lacks the nuance, style, or originality that a human author can provide. Additionally, there is a risk of plagiarism if ChatGPT is not properly trained or if it draws on sources without proper attribution. Therefore, it is important to carefully evaluate and edit any content generated by ChatGPT to ensure that it meets high standards of quality and originality. | **ChatGPT can aid in the editing process, but human expertise, creativity, and critical thinking are essential. Careful evaluation and editing are needed to ensure quality, originality, and avoid plagiarism.** |
| 6 | **Role of CHATGPT so far in neurological disorders related clinical practice and research.** | While there is limited specific information on the role of ChatGPT in neurological disorders related to clinical practice and research, it is worth noting that the integration of artificial intelligence, natural language processing, and GPT technologies in psychiatry shows promise in transforming the diagnosis, treatment, and monitoring of psychiatric disorders. These technologies have the potential to assist psychiatrists in making more accurate diagnoses, developing personalized treatment | **While there is no specific information on ChatGPT's role in neurological disorders, AI technologies** |
|  |  | plans, enhancing the accuracy and efficiency of clinical trials, and increasing access to mental health care through the use of chatbots and virtual assistants. Although there may not be explicit examples or studies on ChatGPT's application in neurological disorders, similar applications could potentially be developed in the future. | **show promise in revolutionizing psychiatric diagnosis, treatment, and accessibility to mental health care.** |
| 7 | **Effectiveness and efficiency of Chat GPT in medical research and clinical settings** | Upon review, I couldn't find specific information on the effectiveness and efficiency of ChatGPT in medical research and clinical settings. The provided pages discuss the potential applications of AI and natural language processing in medicine, including psychiatry, but do not offer specific studies or data on the use of ChatGPT in these settings.<br>It is important to emphasize that while AI tools like ChatGPT show promise in improving medical research and clinical practice, they should not be seen as a substitute for human expertise. Instead, they should be utilized alongside established best practices and ethical guidelines. Further research is necessary to assess the effectiveness and efficiency of ChatGPT specifically in medical research and clinical contexts. | **There is no specific information on ChatGPT's effectiveness in medical research and clinical settings, highlighting the need for further research and consideration of ethical guidelines.** |
| 8 | **Potential benefits and limitations of Chat GPT in medical research and clinical applications** | ChatGPT presents potential benefits and limitations in medical research and clinical applications. Some benefits include its assistance in patient questionnaires, interviews, and epidemiological research by streamlining data collection, recording, and analysis. The integration of artificial intelligence, natural language processing, and GPT technologies in psychiatry holds promise for transforming psychiatric diagnosis, treatment, and monitoring. It can aid in accurate diagnoses, personalized treatment design, improved clinical trial efficiency, and increased accessibility to mental health care through chatbots and virtual assistants. ChatGPT can also be useful for tasks like proofreading and manuscript checks. | **ChatGPT has potential benefits in mental health care, data collection, and manuscript checks but has limitations in providing comprehensive diagnoses and** |
|  |  | However, there are limitations to consider. ChatGPT cannot update its training data in real-time and may provide general or vague answers in medical-related conversations. Its diagnoses may not be comprehensive or sufficient, indicating that it cannot independently handle the complexities of clinical practice. It cannot replace the specialized knowledge, innovative ideas, and opinions that human input provides. Ethical guidelines and further exploration of research ethics are necessary for the responsible use of AI in health research. The quality of AI-generated content requires careful evaluation as it cannot substitute for human expertise, creativity, and critical thinking. While ChatGPT can comprehend information and connect evidence rapidly, it lacks the nuance and originality that human authors provide. Moreover, there is a risk of plagiarism if ChatGPT is not appropriately trained or if it fails to provide proper source attribution. | **replacing human expertise, creativity, and critical thinking. Careful evaluation and ethical guidelines are necessary.** |
| 9 | **The ethical implications of using Chat GPT in medical research and clinical practice** | There are ethical implications of using ChatGPT in medical research and clinical practice. - Almost 97% of the records expressed concerns or possible risks associated with ChatGPT. Ethical issues, such as risk of bias, plagiarism, copyright issues, transparency issues, legal concerns, lack of originality, incorrect responses, limited knowledge, and inaccurate citations were the most common. The quality of the AI-generated content needs careful evaluation as it is not a substitute for human expertise, creativity, and critical thinking. While ChatGPT can rapidly comprehend information and connect evidence, it lacks the nuance or originality that a human author can provide. Additionally, there is a risk of plagiarism if ChatGPT is not properly trained or if it draws on sources without proper attribution. Further exploration of research ethics and the moral use of AI in health research is required to establish guidelines. It is important to consider these ethical implications when using ChatGPT in medical research and clinical practice. Researchers and practitioners should ensure that they are using AI tools like ChatGPT in an ethical manner that respects patient privacy and autonomy while also adhering to<br>established best practices for research ethics. Additionally, they should carefully evaluate any content generated by ChatGPT to ensure that it meets high standards of quality and originality. | **Using ChatGPT in medical research and clinical practice raises ethical concerns such as bias, plagiarism, and limited human-like qualities. Adherence to research ethics and careful evaluation are crucial.** |
| 10 | **Identify the gaps in the current research on Chat GPT and suggest areas for further investigation.** | There are gaps in the current research on ChatGPT and areas for further investigation include: 1. The effectiveness and efficiency of ChatGPT in medical research and clinical settings: While there is potential for ChatGPT to be useful in these settings, there is a lack of specific studies or data on its effectiveness and efficiency. 2. The ethical implications of using ChatGPT in medical research and clinical practice: While there are concerns about ethical issues associated with ChatGPT, further exploration of research ethics and the moral use of AI in health research is required to establish guidelines. 3. The limitations of ChatGPT: While ChatGPT has potential benefits, it also has limitations such as its inability to update its training data in real-time and provide comprehensive diagnoses. Further investigation into these limitations could help improve the technology. 4. The quality of AI-generated content: There is a need for careful evaluation of the quality of AI-generated content as it lacks the nuance or originality that a human author can provide. Further investigation into how to improve the quality of AI-generated content could help make it more useful in medical research and clinical practice. 5. The role of ChatGPT in neurological disorders related clinical practice and research: While there is discussion about the potential applications of AI and natural language processing in medicine, including psychiatry, there is no specific information on the role of ChatGPT in neurological disorders related clinical practice and research. Further investigation into these areas could help improve our understanding of how to best use ChatGPT in medical research and clinical practice while also addressing ethical concerns associated with its use. | **Gaps in current research on ChatGPT include investigating its effectiveness, ethical implications, limitations, content quality, and role in neurological disorders in medical research and clinical practice.** |
| 11 | **Provide insights into the potential future applications of Chat GPT in** | There are potential future applications of ChatGPT in medical research and clinical practice, including: 1. Clinical decision support: ChatGPT could assist in clinical decision-making by providing evidence-based recommendations and identifying patterns and correlations in large | **Potential future applications of ChatGPT in medical research** |
|  | **medical research and clinical practice** | datasets. 2. Clinical trial recruitment: ChatGPT could help identify eligible patients for clinical trials by analyzing electronic health records and other data sources. 3. Clinical data management: ChatGPT could help manage large amounts of clinical data by extracting relevant information from electronic health records and other sources. 4. Research support: ChatGPT could assist researchers in identifying relevant literature, generating hypotheses, and analyzing data. 5. Patient education: ChatGPT could provide personalized education to patients about their conditions, treatments, and medications. 6. Automation of tasks: ChatGPT could automate tasks such as data analysis and image interpretation, freeing up time for healthcare professionals to focus on patient care. 7. Drug discovery: ChatGPT could assist in drug discovery by recognizing, classifying, and describing chemical formulas or molecular structures. 8. Disease prediction, diagnosis, and assessment of therapeutic targets: ChatGPT could help predict diseases based on patient data, diagnose diseases based on symptoms or test results, and assess therapeutic targets for drug development. Overall, the potential future applications of ChatGPT in medical research and clinical practice are vast. However, it is important to address ethical concerns associated with its use while also ensuring that it is used effectively to improve patient outcomes. | **and clinical practice include clinical decision support, trial recruitment, data management, research support, and patient education. Ethical considerations and effective utilization are crucial.** |
| 12 | **Recommendations for researchers, clinicians, and policymakers on the use of ChatGPT in medical research and clinical practice** | 1. Researchers should be transparent about the use of ChatGPT in scientific publications and ensure that the generated text is accurate and reliable.<br>2. Clinicians should use ChatGPT with caution and recognize its limitations in providing reliable information. It can be useful for tasks like proofreading and manuscript checks, but cannot replace the specialized knowledge, innovative ideas, and opinions that human input provides.<br>3. Policymakers should establish regulations and control mechanisms to address ethical utilization of ChatGPT and similar tools in medicine. This includes guidelines for transparency in disclosing the use of AI tools like ChatGPT in scientific publications. | **Researchers, clinicians, and policymakers should ensure transparency, evaluate limitations, establish regulations, and address ethical concerns related** |
|  |  | 4. Researchers should evaluate and monitor the use of ChatGPT to ensure that it is being used ethically and responsibly in medical publishing and research.<br>5. Clinicians should be aware of the potential biases in the training data used to develop ChatGPT and should take steps to mitigate these biases when using the tool.<br>6. Policymakers should consider the potential impact of ChatGPT on clinical and translational medicine and establish guidelines for its use in these areas.<br>7. Researchers, clinicians, and policymakers should work together to address the ethical dilemmas associated with the use of ChatGPT in medical research and clinical practice. | **to ChatGPT in medical applications.** |

## Discussion

We looked into two main uses of ChatGPT: in healthcare settings and for medical writing and research. We studied 118 articles - most were opinion pieces, commentaries, and reviews. Another group, Ruksakulpiwat et al, also did a similar study. They analyzed six articles out of 114 that met their criteria. These articles covered a variety of ways to use ChatGPT, such as finding new drugs, writing literature reviews, improving medical reports, providing medical info, bettering research methods, analyzing data, and personalizing medicine.^7^

Levin et al, on the other hand, conducted an analysis of the first batch of publications about ChatGPT. They found 42 articles published in 26 journals in the 69 days after ChatGPT was launched. Only one was a research article. The rest were mostly editorials and news pieces. Five publications focused on studies on ChatGPT. There were no articles on its use in Obstetrics and Gynecology.In terms of where these articles were published, Nature was the top journal. Radiology and Lancet Digital Health came next. The articles mostly discussed the quality of ChatGPT’s scientific writing, its features, and its performance. Some also talked about who should get credit for the work and ethical concerns. Interestingly, when comparing the articles that described a study to the others, the average impact factor (a measure of the influence of a journal) was significantly lower for the study articles.^8^

In our review, we identified several potential advantages of using ChatGPT in the medical field. It appears to enhance productivity and expedite research workflows by aiding in data organization, assisting in the selection of trial candidates, and supporting overall research activities. Furthermore, ChatGPT’s capacity to review manuscripts and contribute to editing may potentiate the efficiency of academic publishing. Beyond the scope of research, it could also prove beneficial for patient education, fostering scientific exploration, and shaping clinical decision-making. However, we also need to consider certain limitations and ethical concerns associated with the use of ChatGPT. The model, as sophisticated as it is, lacks the capability to offer comprehensive diagnoses and cannot replace the human qualities inherent to medical practice. Ethical issues also arise, specifically in relation to potential biases in the machine learning model and potential breaches of privacy. Moreover, while ChatGPT can process and generate information, it might not exhibit the level of originality, creativity, and critical thinking that are often required in the medical field. However, the use of ChatGPT in producing scholarly articles is raising questions in the academic publishing. While these tools can greatly enhance the clarity and fluency of written material, it is crucial that human oversight is maintained throughout the process. This is because AI can potentially produce content that is authoritative-sounding, yet it might be inaccurate, incomplete, or biased. Incorrect GPT-4 responses, known as “hallucinations,” can be harmful, particularly in the field of medicine. Therefore, it is essential to check or validate GPT-4’s output. ChatGPT can generate references to made-up research publications. Therefore, authors must thoroughly check and modify the output of these tools. Furthermore, it is not appropriate to recognize AI or AI-assisted tools as authors or co-authors in the by-line of publications. Instead, their use should be transparently acknowledged within the manuscript. For example, according to Elsevier’s policy on AI for authors, the responsibility and accountability for the work ultimately still lie with the human authors, despite any technological assistance they may have received.**9-11**

In conclusion, ChatGPT has a great potential. Its full potentials are still evolving. ChatGPT as a source of information can not be trusted, many ethical issues are associated with it. Certainly, ChatGPT can be credited with authorship. However, ChatGPT is certainly a good clinical assistant. ChatGPT is nowhere near to replace human brain.

## Supporting information

Supplementary item-1

## Data Availability

All data produced in the present work are contained in the manuscript.

## Acknowledgment

The concept, data collection analysis, writing, and reporting of this article were solely done by authors. ChatGPT was extensively utilized as mentioned in the methods section.

## Human/Animal Studies informed consent statement

No human or animal subjects were involved.

## References

1. OpenAI. (2023). Conversational AI Model for Medical Inquiries. ChatGPT. https://www.openai.com/chatgpt.

2. Lee P, Bubeck S, Petro J. Benefits, Limits, and Risks of GPT-4 as an AI Chatbot for Medicine. N Engl J Med. 2023 Mar 30;388(13):1233–1239. doi: 10.1056/NEJMsr2214184.

3. Bommasani R, Liang P, Lee T. Holistic Evaluation of Language Models. Ann N Y Acad Sci. 2023 May 25. doi: 10.1111/nyas.15007. Epub ahead of print.

4. Waisberg E, Ong J, Masalkhi M, Kamran SA, Zaman N, Sarker P, Lee AG, Tavakkoli A. GPT-4: a new era of artificial intelligence in medicine. Ir J Med Sci. 2023 Apr 19. doi: 10.1007/s11845-023-03377-8. Epub ahead of print.

5. Ong CWM, Blackbourn HD, Migliori GB. GPT-4, artificial intelligence and implications for publishing. Int J Tuberc Lung Dis. 2023 Jun 1;27(6):425–426. doi: 10.5588/ijtld.23.0143.

6. Garg RK, Paliwal V, Kar SK, Urs VL, Chaudhary SK. Exploring the Role of Chat GPT in patient care (diagnosis and Treatment) and medical research: A Systematic Review. PROSPERO 2023 CRD42023415845 Available from: https://www.crd.york.ac.uk/prospero/display_record.php?ID=CRD42023415845.

7. Ruksakulpiwat S, Kumar A, Ajibade A. Using ChatGPT in Medical Research: Current Status and Future Directions. J Multidiscip Healthc. 2023 May 30;16:1513–1520. doi: 10.2147/JMDH.S413470.

8. Levin G, Brezinov Y, Meyer R. Exploring the use of ChatGPT in OBGYN: a bibliometric analysis of the first ChatGPT-related publications. Arch Gynecol Obstet. 2023 May 24. doi: 10.1007/s00404-023-07081-x. Epub ahead of print.

9. Anonymous. Tools such as ChatGPT threaten transparent science; here are our ground rules for their use. Nature. 2023 Jan;613(7945):612. doi: 10.1038/d41586-023-00191-1.

10. Hill-Yardin EL, Hutchinson MR, Laycock R, Spencer SJ. A Chat(GPT) about the future of scientific publishing. Brain Behav Immun. 2023 May;110:152–154. doi: 10.1016/j.bbi.2023.02.022. Epub 2023 Mar 1.

11. Thorp HH. ChatGPT is fun, but not an author. Science. 2023 Jan 27;379(6630):313. doi: 10.1126/science.adg7879. Epub 2023 Jan 26.

## References

1. Ali MJ, Djalilian A. Readership Awareness Series - Paper 4: Chatbots and ChatGPT - Ethical Considerations in Scientific Publications. Semin Ophthalmol. 2023 Mar 21:1–2. doi: 10.1080/08820538.2023.2193444. Epub ahead of print.

2. Ali SR, Dobbs TD, Hutchings HA, Whitaker IS. Using ChatGPT to write patient clinic letters. Lancet Digit Health. 2023 Apr;5(4):e179–e181. doi: 10.1016/S2589-7500(23)00048-1. Epub 2023 Mar 7.

3. Alser M, Waisberg E. Concerns with the Usage of ChatGPT in Academia and Medicine: A Viewpoint. American Journal of Medicine Open 2023;9. https://doi.org/10.1016/j.ajmo.2023.100036.

4. Anderson N, Belavy DL, Perle SM, Hendricks S, Hespanhol L, Verhagen E, Memon AR. AI did not write this manuscript, or did it? Can we trick the AI text detector into generated texts? The potential future of ChatGPT and AI in Sports & Exercise Medicine manuscript generation. BMJ Open Sport Exerc Med. 2023 Feb 16;9(1):e001568. doi: 10.1136/bmjsem-2023-001568.

5. Arun Babu T, Sharmila V. Using artificial intelligence chatbots like ‘ChatGPT’ to draft articles for medical journals - Advantages, limitations, ethical concerns and way forward. Eur J Obstet Gynecol Reprod Biol. 2023 May 16:S0301–2115(23)00187–2. doi: 10.1016/j.ejogrb.2023.05.008. Epub ahead of print.

6. Asch DA. An Interview with ChatGPT About Health Care. NEJM Catalyst Innovations in Care Delivery 2023;2. https://catalyst.nejm.org/doi/abs/10.1056/CAT.23.0043.

7. Athaluri SA, Manthena SV, Kesapragada VSRKM, Yarlagadda V, Dave T, Duddumpudi RTS. Exploring the Boundaries of Reality: Investigating the Phenomenon of Artificial Intelligence Hallucination in Scientific Writing Through ChatGPT References. Cureus. 2023 Apr 11;15(4):e37432. doi: 10.7759/cureus.37432.

8. Balas M, Ing EB. Conversational AI models for ophthalmic diagnosis: Comparison of ChatGPT and the isabel pro differential diagnosis generator. JFO Open Ophthalmology. 2023 Mar 1;1:100005. https://doi.org/10.1016/j.jfop.2023.100005.

9. Barker FG, Rutka JT. Editorial. Generative artificial intelligence, chatbots, and the Journal of Neurosurgery Publishing Group. J Neurosurg. 2023 Apr 28:1–3. doi: 10.3171/2023.4.JNS23482. Epub ahead of print.

10. Bauchner H. ChatGPT: Not An Author, But A Tool. Health Affairs Forefront. 2023. DOI: 10.1377/forefront.20230511.917632.

11. Baumgartner C. The potential impact of ChatGPT in clinical and translational medicine. Clin Transl Med. 2023 Mar;13(3):e1206. doi: 10.1002/ctm2.1206.

12. Benoit JR. ChatGPT for Clinical Vignette Generation, Revision, and Evaluation. medRxiv 2023.02.04.23285478; doi: https://doi.org/10.1101/2023.02.04.23285478.

13. Bhattacharya K, Bhattacharya AS, Bhattacharya N, Yagnik VD, Garg P, Kumar S. ChatGPT in surgical practice—a New Kid on the Block. Indian Journal of Surgery. 2023 Feb 22:1–4. https://doi.org/10.1007/s12262-023-03727-x.

14. Biswas S. ChatGPT and the Future of Medical Writing. Radiology. 2023 Apr;307(2):e223312. doi: 10.1148/radiol.223312.

15. Boßelmann CM, Leu C, Lal D. Are AI language models such as ChatGPT ready to improve the care of individuals with epilepsy? Epilepsia. 2023 May;64(5):1195–1199. doi: 10.1111/epi.17570.

16. Brainard J. Journals take up arms against AI-written text. Science. 2023 Feb 24;379(6634):740–741. doi: 10.1126/science.adh2762. Epub 2023 Feb 23.

17. Cahan P, Treutlein B. A conversation with ChatGPT on the role of computational systems biology in stem cell research. Stem Cell Reports. 2023 Jan 10;18(1):1–2. doi: 10.1016/j.stemcr.2022.12.009.

18. Nasrallah HA. A ‘guest editorial’… generated by ChatGPT? Current Psychiatry 2023:6. https://cdn.mdedge.com/files/s3fs-public/CP02204006.pdf.

19. Cascella M, Montomoli J, Bellini V, Bignami E. Evaluating the Feasibility of ChatGPT in Healthcare: An Analysis of Multiple Clinical and Research Scenarios. J Med Syst. 2023 Mar 4;47(1):33. doi: 10.1007/s10916-023-01925-4.

20. Chen S, Kann BH, Foote MB, Aerts HJ, Savova GK, Mak RH, Bitterman DS. The utility of ChatGPT for cancer treatment information medRxiv 2023.03.16.23287316; doi: https://doi.org/10.1101/2023.03.16.23287316.

21. Cheng K, Wu H, Li C. ChatGPT/GPT-4: enabling a new era of surgical oncology. Int J Surg. 2023 May 16. doi: 10.1097/JS9.0000000000000451. Epub ahead of print.

22. Chervenak J, Lieman H, Blanco-Breindel M, Jindal S. The promise and peril of using a large language model to obtain clinical information: ChatGPT performs strongly as a fertility counseling tool with limitations. Fertil Steril. 2023 May 20:S0015–0282(23)00522-8. doi: 10.1016/j.fertnstert.2023.05.151. Epub ahead of print.

23. Cifarelli CP, Sheehan JP. Large language model artificial intelligence: the current state and future of ChatGPT in neuro-oncology publishing. J Neurooncol. 2023 May 20. doi: 10.1007/s11060-023-04336-0. Epub ahead of print.

24. Corsello A, Santangelo A. May Artificial Intelligence Influence Future Pediatric Research?-The Case of ChatGPT. Children (Basel). 2023 Apr 21;10(4):757. doi: 10.3390/children10040757.

25. D’Amico RS, White TG, Shah HA, Langer DJ. I Asked a ChatGPT to Write an Editorial About How We Can Incorporate Chatbots Into Neurosurgical Research and Patient Care…. Neurosurgery. 2023 Apr 1;92(4):663–664. doi: 10.1227/neu.0000000000002414. Epub 2023 Feb 9.

26. Darkhabani M, Alrifaai MA, Elsalti A, Dvir YM, Mahroum N. ChatGPT and autoimmunity - A new weapon in the battlefield of knowledge. Autoimmun Rev. 2023 May 19:103360. doi: 10.1016/j.autrev.2023.103360. Epub ahead of print.

27. Dave, M. Plagiarism software now able to detect students using ChatGPT. Br Dent J 234, 642 (2023). https://doi.org/10.1038/s41415-023-5868-8.

28. Dave T, Athaluri SA, Singh S. ChatGPT in medicine: an overview of its applications, advantages, limitations, future prospects, and ethical considerations. Front Artif Intell. 2023 May 4;6:1169595. doi: 10.3389/frai.2023.1169595.

29. Day T. A Preliminary Investigation of Fake Peer-Reviewed Citations and References Generated by ChatGPT. The Professional Geographer. 2023 Mar 23:1–4. DOI: 10.1080/00330124.2023.2190373.

30. de Oliveira RS, Ballestero M. The future of Pediatric Neurosurgery and ChatGPT: an editor’s perspective. Archives of Pediatric Neurosurgery 2023;5. DOI: https://doi.org/10.46900/apn.v5i2.191.

31. De Vito EL. Artificial intelligence and chatGPT. Would you read an artificial author? Medicina 2023;83:329–332.

32. Dergaa I, Chamari K, Zmijewski P, Ben Saad H. From human writing to artificial intelligence generated text: examining the prospects and potential threats of ChatGPT in academic writing. Biol Sport 2023;40:615–622.

33. Donato H, Escada P, Villanueva T. The Transparency of Science with ChatGPT and the Emerging Artificial Intelligence Language Models: Where Should Medical Journals Stand? Acta Med Port. 2023 Mar 1;36(3):147–148. doi: 10.20344/amp.19694. Epub 2023 Feb 9.

34. Dunn C, Hunter J, Steffes W, Whitney Z, Foss M, Mammino J, Leavitt A, Hawkins SD, Dane A, Yungmann M, Nathoo R. Artificial intelligence-derived dermatology case reports are indistinguishable from those written by humans: A single-blinded observer study. J Am Acad Dermatol. 2023 Apr 11:S0190–9622(23)00587-X. doi: 10.1016/j.jaad.2023.04.005. Epub ahead of print.

35. Fatani B. ChatGPT for Future Medical and Dental Research. Cureus. 2023 Apr 8;15(4):e37285. doi: 10.7759/cureus.37285.

36. Galland J. Les chatbots en médecine interne : opportunités et défis à venir [Chatbots and internal medicine: Future opportunities and challenges]. Rev Med Interne. 2023 May;44(5):209–211. French. doi: 10.1016/j.revmed.2023.04.001. Epub 2023 Apr 29.

37. Gandhi Periaysamy A, Satapathy P, Neyazi A, Padhi BK. ChatGPT: roles and boundaries of the new artificial intelligence tool in medical education and health research - correspondence. Ann Med Surg (Lond) 2023;85:1317–1318.

38. Gao CA, Howard FM, Markov NS, Dyer EC, Ramesh S, Luo Y, Pearson AT. Comparing scientific abstracts generated by ChatGPT to original abstracts using an artificial intelligence output detector, plagiarism detector, and blinded human reviewers. bioRxiv 2022.12.23.521610; doi: https://doi.org/10.1101/2022.12.23.521610.

39. Goedde D, Noehl S, Wolf C, Rupert Y, Rimkus L, Ehlers J, Breuckmann F, Sellmann T. ChatGPT in medical literature-a concise review and SWOT analysis. medRxiv 2023.05.06.23289608; doi: https://doi.org/10.1101/2023.05.06.23289608.

40. Gordijn B, Have HT. ChatGPT: evolution or revolution? Med Health Care Philos. 2023 Mar;26(1):1–2. doi: 10.1007/s11019-023-10136-0.

41. Gottlieb M, Kline JA, Schneider AJ, Coates WC. ChatGPT and conversational artificial intelligence: Friend, foe, or future of research? Am J Emerg Med. 2023 May 18;70:81–83. doi: 10.1016/j.ajem.2023.05.018. Epub ahead of print.

42. Graf A, Bernardi RE. ChatGPT in Research: Balancing Ethics, Transparency and Advancement. Neuroscience. 2023 Apr 1;515:71–73. doi: 10.1016/j.neuroscience.2023.02.008. Epub 2023 Feb 21.

43. Graham A. ChatGPT and other AI tools put students at risk of plagiarism allegations, MDU warns. BMJ. 2023 May 17;381:1133. doi: 10.1136/bmj.p1133.

44. Gravel J, D’Amours-Gravel M, Osmanlliu E. Learning to fake it: limited responses and fabricated references provided by ChatGPT for medical questions. medRxiv 2023.03.16.23286914; doi: https://doi.org/10.1101/2023.03.16.23286914.

45. Guo E, Gupta M, Sinha S, Rössler K, Tatagiba M, Akagami R, Al-Mefty O, Sugiyama T, Stieg PE, Pickett GE, de Lotbiniere-Bassett M. neuroGPT-X: Towards an Accountable Expert Opinion Tool for Vestibular Schwannoma. medRxiv 2023.02.25.23286117; doi: https://doi.org/10.1101/2023.02.25.23286117.

46. Gurha P, Ishaq N, Marian AJ. ChatGPT and other artificial intelligence chatbots and biomedical writing. J Cardiovasc Aging. 2023;3(2):20. doi: 10.20517/jca.2023.13. Epub 2023 Mar 31.

47. Haemmerli J, Sveikata L, Nouri A, May A, Egervari K, Freyschlag C, Lobrinus JA, Migliorini D, Momjian S, Sanda N, Schaller K. ChatGPT in glioma patient adjuvant therapy decision making: ready to assume the role of a doctor in the tumour board?. medRxiv 2023.03.19.23287452; doi: https://doi.org/10.1101/2023.03.19.23287452.

48. Harskamp RE, De Clercq L. Performance of ChatGPT as an AI-assisted decision support tool in medicine: a proof-of-concept study for interpreting symptoms and management of common cardiac conditions (AMSTELHEART-2). medRxiv 2023.03.25.23285475; doi: https://doi.org/10.1101/2023.03.25.23285475.

49. Hill-Yardin EL, Hutchinson MR, Laycock R, Spencer SJ. A Chat(GPT) about the future of scientific publishing. Brain Behav Immun. 2023 May;110:152–154. doi: 10.1016/j.bbi.2023.02.022. Epub 2023 Mar 1.

50. Hirani R, Farabi B, Marmon S. Experimenting with ChatGPT: Concerns for academic medicine. J Am Acad Dermatol. 2023 May 11:S0190–9622(23)00747–8. doi: 10.1016/j.jaad.2023.04.045. Epub ahead of print.

51. Homolak J. Opportunities and risks of ChatGPT in medicine, science, and academic publishing: a modern Promethean dilemma. Croat Med J. 2023 Feb 28;64(1):1–3. doi: 10.3325/cmj.2023.64.1.

52. Hosseini M, Gao CA, Liebovitz DM, Carvalho AM, Ahmad FS, Luo Y, MacDonald N, Holmes KL, Kho A. An exploratory survey about using ChatGPT in education, healthcare, and research. medRxiv 2023.03.31.23287979; doi: https://doi.org/10.1101/2023.03.31.23287979.

53. Howard A, Hope W, Gerada A. ChatGPT and antimicrobial advice: the end of the consulting infection doctor? Lancet Infect Dis. 2023 Apr;23(4):405–406. doi: 10.1016/S1473-3099(23)00113-5. Epub 2023 Feb 20.

54. Hsu TW, Tsai SJ, Ko CH, Thompson T, Hsu CW, Yang FC, Tsai CK, Tu YK, Yang SN, Tseng PT, Liang CS. Plagiarism, Quality, and Correctness of ChatGPT-Generated vs Human-Written Abstract for Research Paper. Available at SSRN: https://ssrn.com/abstract=4429014 or http://dx.doi.org/10.2139/ssrn.4429014

55. Huang J, Tan M. The role of ChatGPT in scientific communication: writing better scientific review articles. Am J Cancer Res. 2023 Apr 15;13(4):1148–1154.

56. Hurley D. Your AI Program Will Write Your Paper Now: Neurology Editors on Managing Artificial Intelligence Submissions. Neurology Today 2023;23:10-11.

57. Janssen BV, Kazemier G, Besselink MG. The use of ChatGPT and other large language models in surgical science. BJS Open. 2023 Mar 7;7(2):zrad032. doi: 10.1093/bjsopen/zrad03.

58. Johnson SB, King AJ, Warner EL, Aneja S, Kann BH, Bylund CL. Using ChatGPT to evaluate cancer myths and misconceptions: artificial intelligence and cancer information. JNCI Cancer Spectr. 2023 Mar 1;7(2):pkad015. doi: 10.1093/jncics/pkad015.

59. Juhi A, Pipil N, Santra S, Mondal S, Behera JK, Mondal H. The Capability of ChatGPT in Predicting and Explaining Common Drug-Drug Interactions. Cureus. 2023 Mar 17;15(3):e36272. doi: 10.7759/cureus.36272.

60. Kaneda Y. In the Era of Prominent AI, What Role Will Physicians Be Expected to Play? QJM. 2023 May 22:hcad099. doi: 10.1093/qjmed/hcad099. Epub ahead of print.

61. Kim J. Search for Medical Information and Treatment Options for Musculoskeletal Disorders through an Artificial Intelligence Chatbot: Focusing on Shoulder Impingement Syndrome. medRxiv 2022.12.16.22283512; doi: https://doi.org/10.1101/2022.12.16.22283512.

62. Kim SG. Using ChatGPT for language editing in scientific articles. Maxillofac Plast Reconstr Surg. 2023 Mar 8;45(1):13. doi: 10.1186/s40902-023-00381-x.

63. Koo M. The Importance of Proper Use of ChatGPT in Medical Writing. Radiology. 2023 May;307(3):e230312. doi: 10.1148/radiol.230312. Epub 2023 Mar 7.

64. Kumar AH. Analysis of ChatGPT Tool to Assess the Potential of its Utility for Academic Writing in Biomedical Domain. BEMS Reports [Internet]. 2023Feb.2 [cited 2023Jun.10];9(1):24–30. Available from: https://bemsreports.org/index.php/bems/article/view/132.

65. Lee P, Bubeck S, Petro J. Benefits, Limits, and Risks of GPT-4 as an AI Chatbot for Medicine. N Engl J Med. 2023 Mar 30;388(13):1233–1239. doi: 10.1056/NEJMsr2214184.

66. Levin G, Meyer R, Yasmeen A, Young B, Guige PA, Bar-Noy T, Tatar A, Perelstein O, Brezinov Y. ChatGPT-written OBGYN abstracts fool practitioners. Am J Obstet Gynecol MFM. 2023 Apr 29:100993. doi: 10.1016/j.ajogmf.2023.100993. Epub ahead of print.

67. Li H, Moon JT, Purkayastha S, Celi LA, Trivedi H, Gichoya JW. Ethics of large language models in medicine and medical research. Lancet Digit Health. 2023 Jun;5(6):e333–e335. doi: 10.1016/S2589-7500(23)00083-3. Epub 2023 Apr 27.

68. Li S. ChatGPT has made the field of surgery full of opportunities and challenges. Int J Surg. 2023 May 17. doi: 10.1097/JS9.0000000000000454. Epub ahead of print.

69. Liebrenz M, Schleifer R, Buadze A, Bhugra D, Smith A. Generating scholarly content with ChatGPT: ethical challenges for medical publishing. Lancet Digit Health. 2023 Mar;5(3):e105–e106. doi: 10.1016/S2589-7500(23)00019-5. Epub 2023 Feb 6.

70. Lin Z. Modernizing authorship criteria: Challenges from exponential authorship inflation and generative artificial intelligence. 2023. (Preprint). https://psyarxiv.com/s6h58.

71. Loh E. ChatGPT and generative AI chatbots: challenges and opportunities for science, medicine and medical leaders. BMJ Lead. 2023 May 2:leader-2023-000797. doi: 10.1136/leader-2023-000797. Epub ahead of print.

72. Maeker E, Maeker-Poquet B. ChatGPT : une solution pour rédiger des revues de littérature en médecine ?, Volume 8035, Issue 135, 06/2023, Pages 137–212, ISSN 1627-4830, http://dx.doi.org/10.1016/j.npg.2023.03.002 (http://www.sciencedirect.com/science/article/pii/S1627-4830(23)00037-5).

73. Marchandot B, Matsushita K, Carmona A, Trimaille A, Morel O. ChatGPT: the next frontier in academic writing for cardiologists or a pandora’s box of ethical dilemmas. Eur Heart J Open. 2023 Feb 13;3(2):oead007. doi: 10.1093/ehjopen/oead007.

74. Martínez-Sellés M, Marina-Breysse M. Current and Future Use of Artificial Intelligence in Electrocardiography. J Cardiovasc Dev Dis. 2023 Apr 17;10(4):175. doi: 10.3390/jcdd10040175.

75. Mehnen L, Gruarin S, Vasileva M, Knapp B. ChatGPT as a medical doctor? A diagnostic accuracy study on common and rare diseases. medRxiv 2023.04.20.23288859; doi: https://doi.org/10.1101/2023.04.20.23288859.

76. Mello MM, Guha N. ChatGPT and Physicians’ Malpractice Risk. JAMA Health Forum. 2023 May 5;4(5):e231938. doi: 10.1001/jamahealthforum.2023.1938.

77. Mese I. The imperative of a radiology AI deployment registry and the potential of ChatGPT. Clin Radiol. 2023 Jul;78(7):554. doi: 10.1016/j.crad.2023.04.001. Epub 2023 Apr 18.

78. Nastasi NJ, Courtright KR, Halpern SD, Weissman GE. Does ChatGPT Provide Appropriate and Equitable Medical Advice? A Vignette-Based, Clinical Evaluation Across Care Contexts. medRxiv 2023.02.25.23286451; doi: https://doi.org/10.1101/2023.02.25.23286451.

79. Nguyen Y, Costedoat-Chalumeau N. Les intelligences artificielles conversationnelles en médecine interne : l’exemple de l’hydroxychloroquine selon ChatGPT [Artificial intelligence and internal medicine: The example of hydroxychloroquine according to ChatGPT]. Rev Med Interne. 2023 May;44(5):218–226. French. doi: 10.1016/j.revmed.2023.03.017.

80. Nógrádi B, Polgár TF, Meszlényi V, Kádár Z, Hertelendy P, Csáti A, et al. Is There Any Room for Generative AI in Neurology and Other Medical Areas?. Available at SSRN: https://ssrn.com/abstract=4372965 or http://dx.doi.org/10.2139/ssrn.4372965.

81. North RA. >Plagiarism Reimagined. Function. 2023; 4(3):zqad014, https://doi.org/10.1093/function/zqad014.

82. Oh N, Choi GS, Lee WY. ChatGPT goes to the operating room: evaluating GPT-4 performance and its potential in surgical education and training in the era of large language models. Ann Surg Treat Res. 2023 May;104(5):269–273. doi: 10.4174/astr.2023.104.5.269. Epub 2023 Apr 28.

83. Okan Ç. AI and Psychiatry: The ChatGPT Perspective. Alpha Psychiatry. 2023 Mar 1;24(2):41–42. doi: 10.5152/alphapsychiatry.2023.010223.

84. Parsa A, Ebrahimzadeh MH. ChatGPT in Medicine; a Disruptive Innovation or Just One Step Forward? Arch Bone Jt Surg. 2023;11(4):225–226. doi: 10.22038/abjs.2023.22042.

85. Patel SB, Lam K, Liebrenz M. ChatGPT: friend or foe. Lancet Digit Health 2023;5:e102. DOI: https://doi.org/10.1016/S2589-7500(23)00023-7.

86. Pourhoseingholi MA, Hatamnejad MR, Solhpour A. Does chatGPT (or any other artificial intelligence language tool) deserve to be included in authorship list? Gastroenterol Hepatol Bed Bench. 2023;16(1):435–437. doi: 10.22037/ghfbb.v16i1.2747.

87. Rao D. The Urgent Need for Healthcare Workforce Upskilling and Ethical Considerations in the Era of AI-Assisted Medicine. Indian J Otolaryngol Head Neck Surg (2023). https://doi.org/10.1007/s12070-023-03755-9.

88. Ray PP, Majumder P. AI Tackles Pandemics: ChatGPT’s Game-Changing Impact on Infectious Disease Control. Ann Biomed Eng. 2023 May 18:1–3. doi: 10.1007/s10439-023-03239-5. Epub ahead of print.

89. Ros-Arlanzón P, Pérez-Sempere A. ChatGPT: una novedosa herramienta de escritura para artículos científicos, pero no un autor (por el momento) [ChatGPT: a novel tool for writing scientific articles, but not an author (for the time being)]. Rev Neurol. 2023 Apr 16;76(8):277. Spanish. doi: 10.33588/rn.7608.2023066.

90. Sabry Abdel-Messih M, Kamel Boulos MN. ChatGPT in Clinical Toxicology. JMIR Med Educ. 2023 Mar 8;9:e46876. doi: 10.2196/46876.

91. Sallam M. The Utility of ChatGPT as an Example of Large Language Models in Healthcare Education, Research and Practice: Systematic Review on the Future Perspectives and Potential Limitations. medRxiv 2023.02.19.23286155; doi: https://doi.org/10.1101/2023.02.19.23286155.

92. Sallam M, Salim NA, Al-Tammemi AB, Barakat M, Fayyad D, Hallit S, Harapan H, Hallit R, Mahafzah A. ChatGPT Output Regarding Compulsory Vaccination and COVID-19 Vaccine Conspiracy: A Descriptive Study at the Outset of a Paradigm Shift in Online Search for Information. Cureus. 2023 Feb 15;15(2):e35029. doi: 10.7759/cureus.35029.

93. Salvagno M, Taccone FS, Gerli AG. Can artificial intelligence help for scientific writing? Crit Care. 2023 Feb 25;27(1):75. doi: 10.1186/s13054-023-04380-2. Erratum in: Crit Care. 2023 Mar 8;27(1):99.

94. Sanmarchi F, Bucci A, Nuzzolese AG. et al. A step-by-step researcher’s guide to the use of an AI-based transformer in epidemiology: an exploratory analysis of ChatGPT using the STROBE checklist for observational studies. J Public Health (Berl.) (2023). https://doi.org/10.1007/s10389-023-01936-y

95. Sarink MJ, Bakker IL, Anas AA, Yusuf E. A study on the performance of ChatGPT in infectious diseases clinical consultation. Clin Microbiol Infect. 2023 May 18:S1198–743X(23)00241-0. doi: 10.1016/j.cmi.2023.05.017. Epub ahead of print.

96. Schulte B. Capacity of ChatGPT to Identify Guideline-Based Treatments for Advanced Solid Tumors. Cureus. 2023 Apr 21;15(4):e37938. doi: 10.7759/cureus.37938.

97. Singh OP. Artificial intelligence in the era of ChatGPT - Opportunities and challenges in mental health care. Indian J Psychiatry. 2023 Mar;65(3):297–298. doi: 10.4103/indianjpsychiatry.indianjpsychiatry_112_23.

98. Singh S, Djalilian A, Ali MJ. ChatGPT and Ophthalmology: Exploring Its Potential with Discharge Summaries and Operative Notes. Semin Ophthalmol. 2023 May 3:1–5. doi: 10.1080/08820538.2023.2209166. Epub ahead of print.

99. Tang L, Sun Z, Idnay B, Nestor JG, Soroush A, Elias PA, Xu Z, Ding Y, Durrett G, Rousseau J, Weng C, Peng Y. Evaluating Large Language Models on Medical Evidence Summarization. medRxiv 2023.04.22.23288967; doi: https://doi.org/10.1101/2023.04.22.23288967.

100. Temsah O, Khan SA, Chaiah Y, Senjab A, Alhasan K, Jamal A, Aljamaan F, Malki KH, Halwani R, Al-Tawfiq JA, Temsah MH, Al-Eyadhy A. Overview of Early ChatGPT’s Presence in Medical Literature: Insights From a Hybrid Literature Review by ChatGPT and Human Experts. Cureus. 2023 Apr 8;15(4):e37281. doi: 10.7759/cureus.37281.

101. Thorp HH. ChatGPT is fun, but not an author. Science. 2023 Jan 27;379(6630):313. doi: 10.1126/science.adg7879. Epub 2023 Jan 26.

102. Haq ZU, Naeem H, Naeem A, Iqbal F, Zaeem D. Comparing human and artificial intelligence in writing for health journals: an exploratory study. medRxiv 2023.02.22.23286322; doi: https://doi.org/10.1101/2023.02.22.23286322.

103. Uprety D, Zhu D, West HJ. ChatGPT-A promising generative AI tool and its implications for cancer care. Cancer. 2023 May 14. doi: 10.1002/cncr.34827. Epub ahead of print.

104. Uz C, Umay E. “Dr ChatGPT“: Is it a reliable and useful source for common rheumatic diseases? Int J Rheum Dis. 2023 May 23. doi: 10.1111/1756-185X.14749. Epub ahead of print.

105. van Dis EAM, Bollen J, Zuidema W, van Rooij R, Bockting CL. ChatGPT: five priorities for research. Nature. 2023 Feb;614(7947):224-226. doi: 10.1038/d41586-023-00288-7.

106. Waisberg E, Ong J, Masalkhi M, Kamran SA, Zaman N, Sarker P, Lee AG, Tavakkoli A. GPT-4: a new era of artificial intelligence in medicine. Ir J Med Sci. 2023 Apr 19. doi: 10.1007/s11845-023-03377-8. Epub ahead of print.

107. Wen J, Wang W. The future of ChatGPT in academic research and publishing: A commentary for clinical and translational medicine. Clin Transl Med. 2023 Mar;13(3):e1207. doi: 10.1002/ctm2.1207.

108. Xue VW, Lei P, Cho WC. The potential impact of ChatGPT in clinical and translational medicine. Clin Transl Med. 2023 Mar;13(3):e1216. doi: 10.1002/ctm2.1216.

109. Yadava OP. ChatGPT—a foe or an ally?. Indian J Thorac Cardiovasc Surg 2023;39:217–21. https://doi.org/10.1007/s12055-023-01507-6.

110. Au Yeung J, Kraljevic Z, Luintel A, Balston A, Idowu E, Dobson RJ, Teo JT. AI chatbots not yet ready for clinical use. Front Digit Health. 2023 Apr 12;5:1161098. doi: 10.3389/fdgth.2023.1161098.

111. Young JN, O’Hagan R, Poplausky D, et al. The utility of ChatGPT in generating patient-facing and clinical responses for melanoma. J Am Acad Dermatol. 2023 May 18:S0190–9622(23)00908-8. doi: 10.1016/j.jaad.2023.05.024.

112. Zheng H, Zhan H. ChatGPT in Scientific Writing: A Cautionary Tale. Am J Med. 2023 Mar 10:S0002–9343(23)00159-6. doi: 10.1016/j.amjmed.2023.02.011. Epub ahead of print.

113. Zhong Y, Chen YJ, Zhou Y, Lyu YA, Yin JJ, Gao YJ. The Artificial intelligence large language models and neuropsychiatry practice and research ethic. Asian J Psychiatr 2023;84:103577.

114. Zhou J, Jia Y, Qiu Y, Lin L. The Potential of Applying Chatgpt to Extract Keywords of Medical Literature in Plastic Surgery. Aesthet Surg J. 2023 May 20:sjad158. doi: 10.1093/asj/sjad158. Epub ahead of print.

115. Zhou Z. Evaluation of ChatGPT’s Capabilities in Medical Report Generation. Cureus 2023;15:e37589.

116. Zhu L, Mou W, Chen R. Can the ChatGPT and other large language models with internet-connected database solve the questions and concerns of patient with prostate cancer and help democratize medical knowledge? J Transl Med. 2023 Apr 19;21(1):269.

117. Zielinski C, Winker M, Aggarwal R, et al. Chatbots, ChatGPT, and Scholarly Manuscripts: WAME Recommendations on ChatGPT and Chatbots in Relation to Scholarly Publications. Open Access Macedonian Journal of Medical Sciences 2023;11:83–86. DOI: 10.3889/oamjms.2023.11502.

118. Zimmerman A. A Ghostwriter for the Masses: ChatGPT and the Future of Writing. Ann Surg Oncol. 2023 Jun;30(6):3170–3173.

